# NSAID use is associated with lower dementia and Alzheimer’s disease prevalence and slower cognitive decline: A retrospective longitudinal analysis of the NACC cohort

**DOI:** 10.64898/2026.06.27.26355593

**Authors:** Carolin Luisa Hoehne, Victor Salinas, Afsaneh Shirani, Olaf Stuve, Barbara Elena Stopschinski

**Affiliations:** Department of Neurology, Charité Universitätsmedizin Berlin, Bonhoefferweg 3, 10117 Berlin, Germany; Rocky Mountain Movement Disorders Center, Englewood, CO, USA; Saint Luke’s Marion Bloch Neuroscience Institute, Kansas City, MO, USA; Department of Neurology, University of Missouri-Kansas City, Kansas City, MO, USA; Department of Neurology, University of Texas Southwestern Medical Center, Dallas, TX, USA; Peter O’Donnell Brain Institute, University of Texas Southwestern Medical Center, Dallas, TX, USA; Neurology Section, Dallas VA Medical Center, 4500 South Lancaster Road, Dallas, TX, USA; Center for Alzheimer’s and Neurodegenerative Diseases, University of Texas Southwestern Medical Center, Dallas, TX, USA

**Keywords:** NSAIDs, Alzheimer‘s disease, Dementia, neurodegeneration, neuroinflammation, neuroprotection

## Abstract

**INTRODUCTION:** Dementia, particularly Alzheimer’s disease (AD), is a major global health challenge, with prevalence projected to reach 150 million cases by 2050. AD is characterized by progressive cognitive decline linked to neuroinflammation and neurodegeneration. Non-steroidal anti-inflammatory drugs (NSAIDs) have been explored as potential neuroprotective agents, particularly diclofenac, which has been proposed to modulate microglial inflammasome signaling. However, prior studies investigating NSAIDs in AD have yielded inconsistent findings. We therefore reexamined the relationship between selected NSAIDs and dementia outcomes in a large longitudinal cohort from the National Alzheimer’s Coordinating Center (NACC).

**METHODS:** We analyzed cross-sectional and longitudinal data from the NACC database collected between 2005 and 2022. Associations between NSAID exposure and dementia, AD, and cognitive trajectories were examined. Propensity score matching was performed to compare NSAID users with matched non-users while adjusting for demographic and clinical confounders. Longitudinal mixed-effects models were used to assess cognitive decline based on Montreal Cognitive Assessment (MoCA) scores.

**RESULTS:** Among 47,165 participants, diclofenac and naproxen use were associated with a lower prevalence of dementia and AD compared with matched non-users, whereas etodolac showed no significant associations. Diclofenac users demonstrated reduced odds of dementia and AD. Naproxen showed similar cross-sectional associations. In longitudinal modeling, diclofenac users had a significantly slower rate of cognitive decline than non-users.

**DISCUSSION:** These findings suggest a compound-specific association between NSAID use and AD, with diclofenac potentially modulating disease progression through anti-inflammatory mechanisms. The observed modulation of longitudinal cognitive decline supports further investigation of inflammatory pathways, including microglial and inflammasome signaling, as therapeutic targets in biomarker-defined AD populations.

## 1. Background

Dementia, including Alzheimer’s disease (AD), remains a leading cause of morbidity in aging populations worldwide [1]. AD accounts for nearly 80% of dementia cases, and is characterized by progressive decline in memory, executive function, and other cognitive domains, ultimately impairing activities of daily living and quality of life [3].

AD is defined by the accumulation of extracellular amyloid-β (Aβ) aggregates and intracellular neurofibrillary tangles composed of tau protein aggregates [4]. Pathological tau aggregates accumulate in a pattern predictive of disease progression, first described by Braak *et al.* [4]. Evidence shows that tau aggregates propagate along a predefined neuronal network from cell to cell, thus inducing tau alteration in neighboring cells [5]. Microglia, resident phagocytic immune cells in the brain, contribute to this prion-like process [6]. Pathological tau and Aβ aggregates stimulate microglia [7–11], which in turn launches downstream pro-inflammatory pathways. For example, activation of the microglial Nod-like receptor family, pyrin domain-containing 3 (NLRP3)-inflammasome results in secretion of interleukin-1β (IL-1β), which can promote progression of AD pathology [7–11]. Although innate immune activation is traditionally viewed as protective, e.g., against invading pathogens, these findings indicate that chronic immune activation can be detrimental in AD. Emerging data further suggest that central nervous system (CNS)-infiltrating peripheral immune cells, including myeloid cells, T and B cells, contribute to disease progression [12–15].

The growing evidence for the prominent role of inflammation in AD sparked an early interest in non-steroidal anti-inflammatory drugs (NSAIDs) as potential neuroprotective agents [16]. NSAIDs act primarily through inhibition of cyclooxygenase (COX) enzymes, reducing prostaglandin-mediated inflammatory signaling, and encompass multiple chemical classes with differing COX selectivity, pharmacokinetics, and safety profiles, among them indole acids (e.g. etodolac), and arylacetic acids (e.g. diclofenac) and propionic acid derivatives (e.g., naproxen)[17]. For certain NSAIDs, additional mechanistic pathways have been postulated. For example, some studies suggest that diclofenac modulates neuroinflammation by inhibiting the NLRP3 and IL-1β pathways, with a potential beneficial effect on AD pathology [6,18].

Epidemiological studies from the late 1990s suggested that NSAID use might be associated with a reduced AD risk [19–21]. However, other studies failed to demonstrate a consistent therapeutic benefit of specific NSAIDs for preventing or slowing AD progression [22–24]. These discrepancies can be attributed in part to the timing of intervention, the lack of disease staging, heterogeneity in diagnosis, and the absence of subgroup analyses. With the recent availability of anti-amyloid therapies and promising preclinical evidence for other therapeutic approaches (such as genetic modulation of tau expression, anti-tau antibodies, and modulation of selected inflammatory pathways), research has shifted away from further evaluation of NSAIDs in AD, despite their clinical availability.

In recent years, advances in biomarker-driven diagnostics, including cerebrospinal fluid (CSF) and imaging-based measures of amyloid and tau pathology, have redefined how AD is diagnosed and classified [25]. These diagnostic tools have enabled distinction among pathophysiological subtypes and targeted investigation of disease-modifying factors and treatment outcomes.

In this study, we leveraged a longitudinal cohort from the National Alzheimer’s Coordinating Center (NACC) database, which includes detailed NSAID exposure, diagnostic classification, and CSF biomarker data, to reexamine the effects of NSAIDs, particularly diclofenac, naproxen, and etodolac, on the prevalence and progression of dementia and AD.

## 2. Methods

### 2.1. Study design and objective

This retrospective cohort study was conducted using a dataset provided by the NACC at University of Washington (Seattle, WA, USA). The study’s primary objective was to determine whether diclofenac use is associated with a reduced prevalence of dementia and AD compared to naproxen, etodolac, and no NSAID use. The secondary objective was to assess whether users of diclofenac demonstrate better cognitive performance, as measured by the MoCA. Finally, as an exploratory aim, we examined whether NSAID exposure influenced longitudinal cognitive trajectories.

### 2.2. Dataset

NACC curates a large, centralized relational database of standardized clinical research data contributed by National Institute on Aging (NIA)–funded Alzheimer’s Disease Research Centers (ADRC) across the United States. To harmonize data collection across the 29 ADRCs, the NIA ADRC Clinical Task Force developed the Uniform Data Set (UDS), a standardized clinical dataset including demographic, clinical, and cognitive assessments. The dataset request was submitted to NACC on 02/20/2023, and the requested data files were received on 03/06/2023. The provided data included all visits for all participants as of the December 2022 data freeze and contained patient data collected during visits conducted between June 2005 and November 2022.

### 2.3. Participants

Each participant had a unique Study ID with multiple visits. For cross-sectional analyses, participants were collapsed to the most recent visit and MoCA score. The presence of any comorbid conditions was retained if indicated at any visit, and the use of medications was included if listed in the drug inventory at any visit. A carefully selected list of NACC variables was included in our analysis (***Supplementary Table 1***). NSAID exposure was classified as (1) none; (2) exclusive use of diclofenac, naproxen, or etodolac; or (3) multiple use if two or more NSAIDs were at least once recorded. The duration of NSAID was not recorded in the NACC datasets and is therefore not included in our study.

Cognitive status was defined as follows: participants who met NACC criteria for all-cause dementia were classified as demented. In contrast, those with normal cognition or with cognitive/behavioral impairment not meeting dementia criteria were classified as non-demented. Dementia criteria varied across NACC UDS versions: earlier versions lacked standardized definitions, while version 3 included formal criteria (Form D1 see ***Supplementary Figure 1***).

AD was defined as cognitive impairment with an etiologic AD diagnosis following the NINCDS-ADRDA (UDS versions 1-2) and the National Institute on Aging-Alzheimer’s Association (NIA-AA) guidelines (UDS versions 3-3.2) [27,28]. In the cross-sectional analysis, participants with cognitive impairment due to other causes were excluded, and those with normal cognition served as the reference group.

### 2.4. Statistical Analysis

We conducted all analyses using R within the RStudio v2024.12.1+563 environment (Posit PBC, Boston, MA, USA). The R analysis script and the complete reproducible workflow are provided as a supplementary file.

Continuous variables were summarized as median (interquartile range) and compared using the Kruskal–Wallis tests while categorical variables were compared using Pearson’s chi-squared test.

Propensity score matching without replacement was performed using the *MatchIt* package in R, separately comparing users of diclofenac, naproxen, and etodolac with participants reporting no NSAID use. Propensity scores were estimated by logistic regression, including age, sex, education, race, and prespecified comorbidities (traumatic brain injury, depression, bipolar disorder, schizophrenia, anxiety, post-traumatic stress disorder, cancer, diabetes, congestive heart failure, hypertension, hypercholesterolemia, vitamin B12 deficiency, thyroid disease, arthritis, sleep apnea). Post-matching outcomes for each NSAID user and non-user group were assessed using chi-squared tests (cognitive status, AD), Wilcoxon tests (MoCA), and unadjusted logistic regression models, reporting odds ratios with 95% confidence intervals.

Longitudinal MoCA trajectories were analyzed using linear mixed-effects models with random intercepts and slopes, including time, NSAID group, and their interaction to estimate annualized cognitive change.

## 3. Results

### 3.1. Study population

Table 1 summarizes the demographic and clinical characteristics of the 47,165 participants included from the NACC database. The median age was 75 years (interquartile range [IQR]: 68-82), and 57% were female. Most participants identified as White (79%), followed by Black or African American (13%). The median education-adjusted MoCA score was 24 (IQR: 19-27). Overall, 43% of participants were classified as demented, and 44% carried a presumptive (i.e., not biomarker-based) diagnosis of AD. CSF biomarker data were collected for a subset of participants. CSF tau (total tau or phospho-tau) data were available for a subset of participants (n=1,434), of whom 46.5% exhibited elevated tau levels (positive). CSF Aβ data were available for 1,523 participants, with 50.9% classified as low amyloid (positive). When CSF biomarker profiles were considered jointly, the most common patterns were concordant normal tau and normal amyloid (42%) followed by elevated tau and low amyloid (39%), whereas isolated low amyloid or isolated elevated tau were less common. Regarding NSAID use, naproxen was the most frequently reported agent (5.8%), followed by diclofenac (1.9%).

**Table 1.** Demographic and clinical characteristics of NACC cohort. The diagnosis of Alzheimer’s disease (AD) based on the NACC variables was categorized into normal cognition, presumptive AD diagnosis, and no diagnosis. MoCA, Montreal Cognitive Assessment; NSAID, Non-steroidal anti-inflammatory drugs. Median (Interquartile Range (IQR); n (%). Additional details are discussed in the text.

| **Variable** | **N = 47,165** |
| --- | --- |
| **Age in Years** | 75 (68-82) |
| **Sex** |  |
| Male | 20,263 (43%) |
| Female | 26,902 (57%) |
| **Race** |  |
| White | 37,297 (79%) |
| Black or African American | 5,996 (13%) |
| American Indian or Alaska Native | 288 (0.6%) |
| Native Hawaiian or Pacific Islander | 36 (<0.1%) |
| Asian | 1,265 (2.7%) |
| Multiracial | 1,459 (3.1%) |
| Unknown | 824 (1.7%) |
| **Education (years)** | 16 (12-18) |
| **MoCA score (Education-corrected)** | 24 (19-27) |
| **Cognitive Status (demented)** | 20,053 (43%) |
| **Evidence of Alzheimer's disease** |  |
| Normal cognition | 9,696 (21%) |
| Diagnosis (presumptive) | 20,644 (44%) |
| No Diagnosis | 16,825 (36%) |
| **Elevated Tau in Cerebrospinal Fluid** |  |
| Positive | 667 (47%) |
| Negative | 767 (53%) |
| **Low Amyloid Cerebrospinal Fluid** |  |
| Positive | 775 (51%) |
| Negative | 748 (49%) |
| **Cerebrospinal Fluid- Status** |  |
| Elevated Tau and low Amyloid | 543 (39%) |
| Normal Tau and normal Amyloid | 583 (42%) |
| Only low Amyloid | 151 (11%) |
| Only elevated Tau | 98 (7.1%) |
| **NSAID Use** |  |
| Naproxen | 2,715 (5.8%) |
| Diclofenac | 879 (1.9%) |
| Etodolac | 151 (0.3%) |
| Multiple NSAID | 174 (0.4%) |
| No NSAID | 43,246 (92%) |

### 3.2. NSAID use and prevalence of dementia and Alzheimer’s disease

We hypothesized that, specifically, the use of diclofenac alone, and no other NSAIDs, would be linked to a lower prevalence of dementia and AD compared to no NSAID use. This hypothesis was motivated by our prior work suggesting a diclofenac-specific neuroprotective effect [18]. In two U.S. veteran cohorts, Stuve et al. reported significantly lower AD incidence among diclofenac users than among those using naproxen or etodolac [18].

To examine this, we grouped participants by reported medication use: naproxen, diclofenac, etodolac, or no NSAID (***Table 2***). Significant differences were found across these groups in age, sex, race, MoCA scores, and the prevalence of dementia and AD (all P values < .001). Participants not using NSAIDs were generally younger, had lower MoCA scores, and a higher prevalence of dementia and AD, compared to NSAID users of any kind. These differences led us to apply propensity score matching [29] to adjust for potential confounding factors.

**Table 2.**
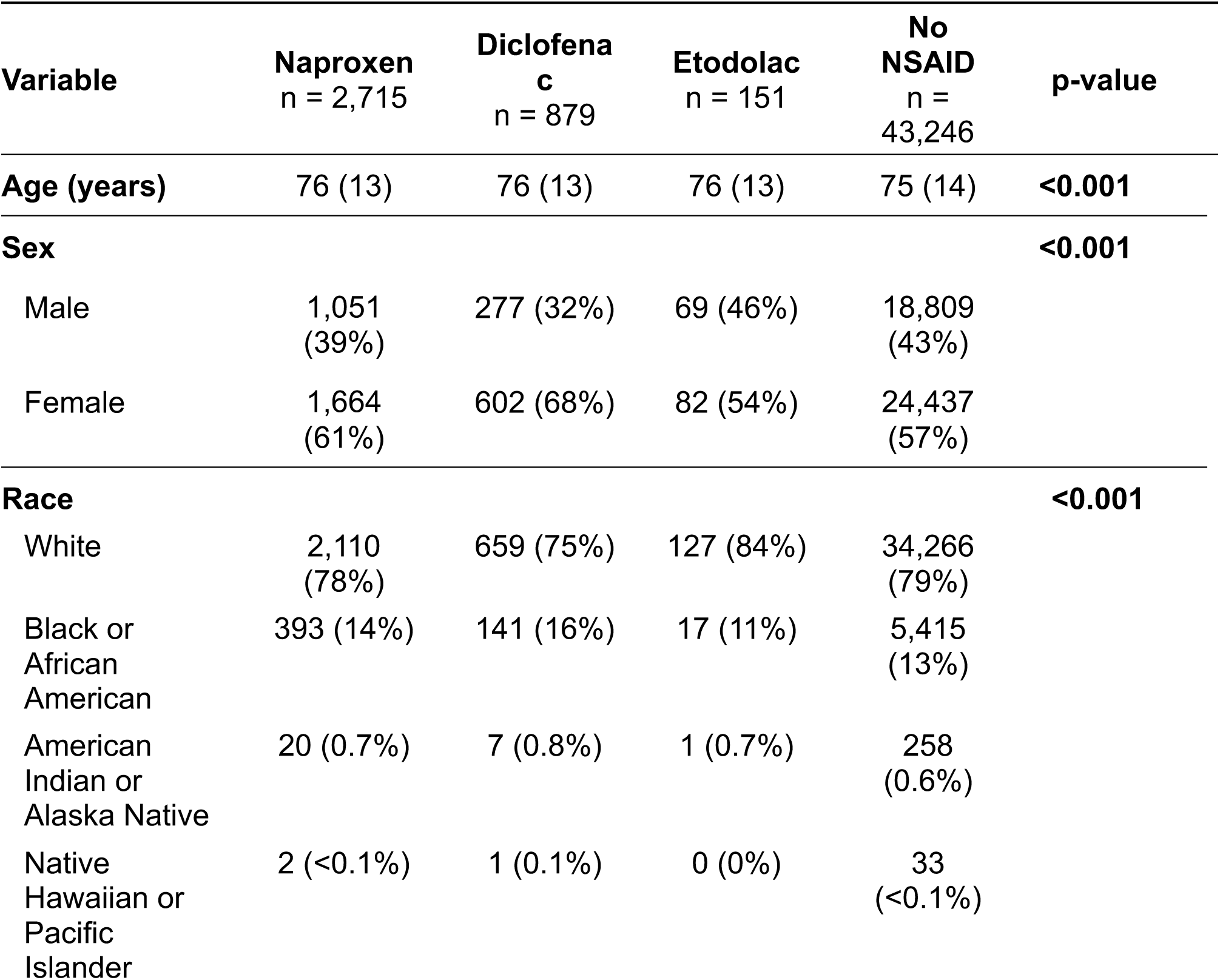
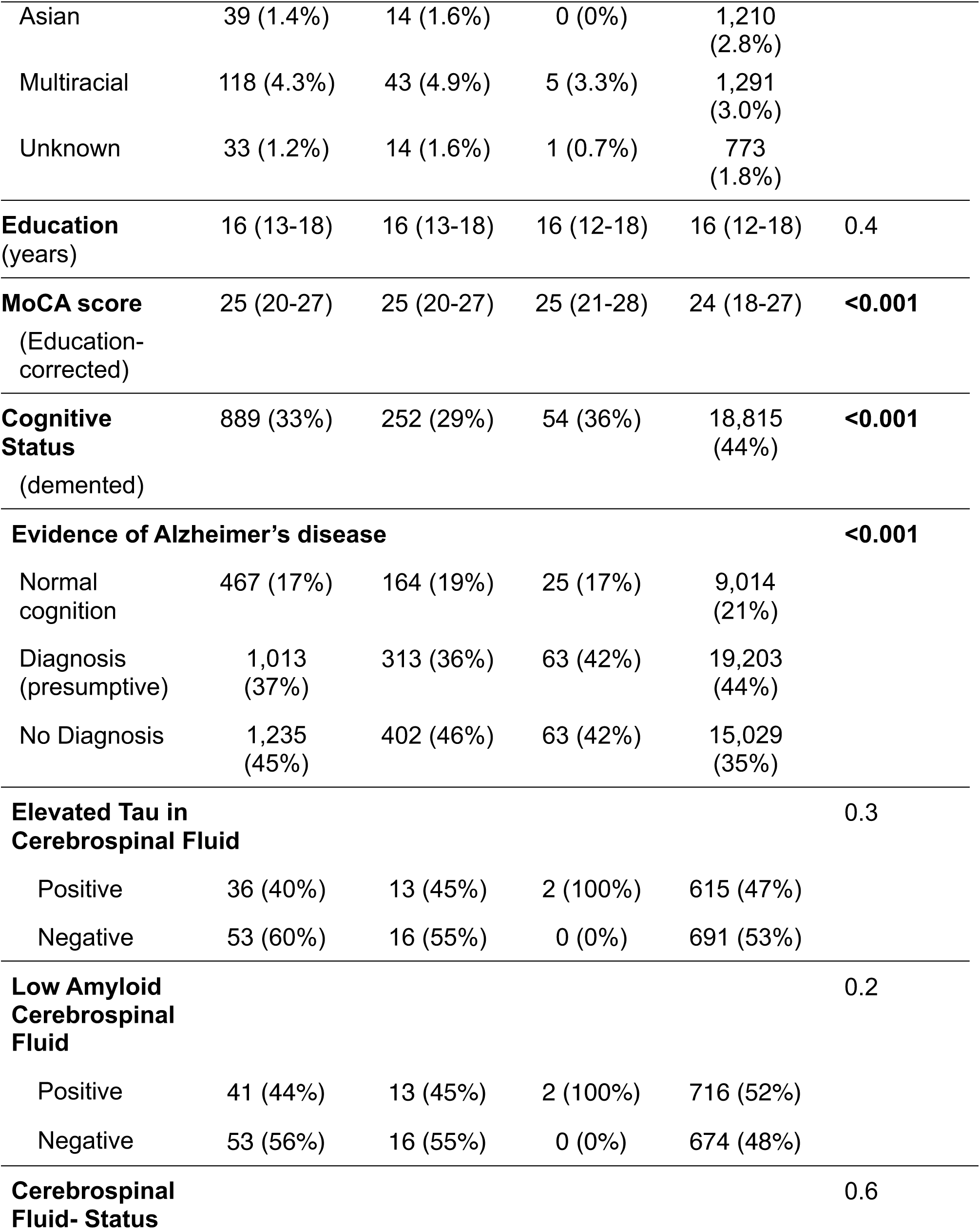
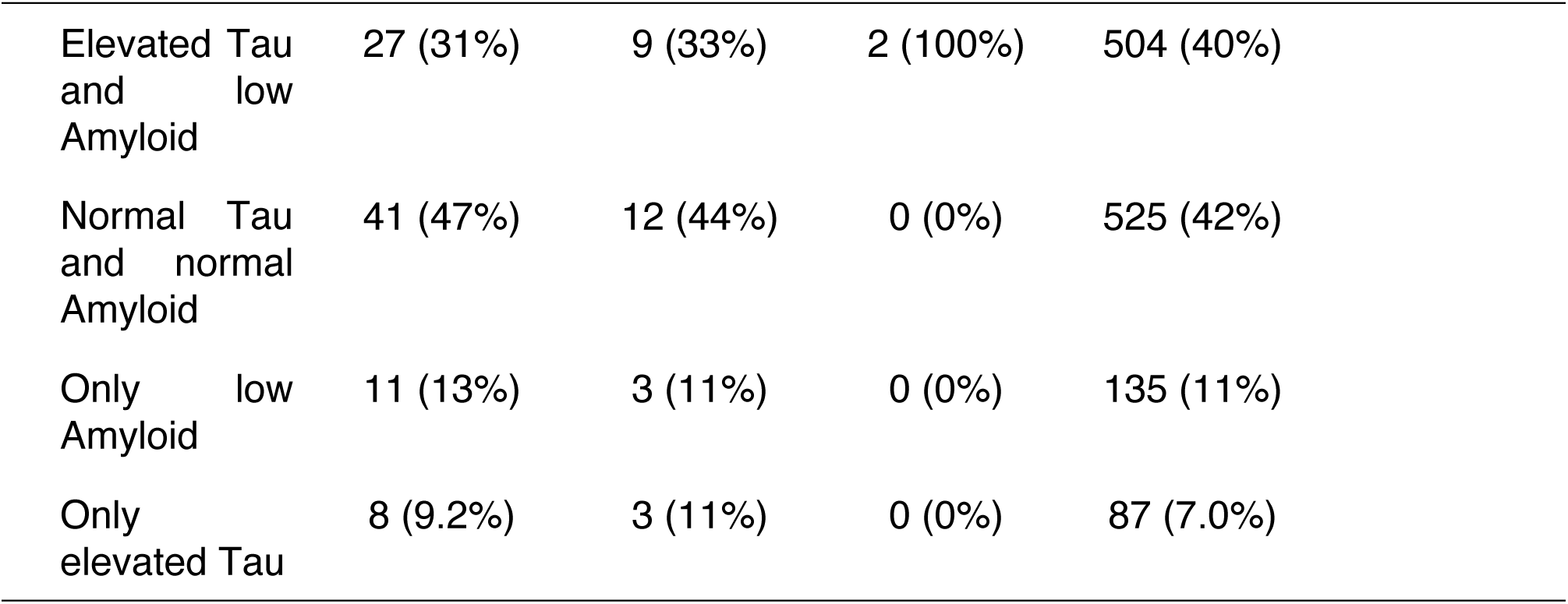
Demographic and clinical characteristics of participants in the NACC cohort by NSAID use. The cohort was grouped into those who used a single NSAID (Diclofenac, Naproxen, Etodolac) or no NSAID. Participants who reported use of multiple NSAIDs (more than two different NSAIDs) (N = 174) were excluded from the analysis. Continuous variables are summarized as median and interquartile range (IQR), and categorical variables as counts and percentages. Comparisons between groups were conducted using the Kruskal–Wallis test for continuous variables and chi-square tests for categorical variables—AD, Alzheimer’s disease; MoCA, Montreal Cognitive Assessment; NSAID, Non-steroidal anti-inflammatory drugs.

| **Variable** | **Naproxen**<br>**n = 2,715** | **Diclofenac**<br>**n = 879** | **Etodolac**<br>**n = 151** | **No**<br>**NSAID**<br>**n =**<br>**43,246** | **p-value** |
| --- | --- | --- | --- | --- | --- |
| **Age (years)** | 76 (13) | 76 (13) | 76 (13) | 75 (14) | **<0.001** |
| **Sex** |  |  |  |  | **<0.001** |
| Male | 1,051<br>(39%) | 277 (32%) | 69 (46%) | 18,809<br>(43%) |  |
| Female | 1,664<br>(61%) | 602 (68%) | 82 (54%) | 24,437<br>(57%) |  |
| **Race** |  |  |  |  | **<0.001** |
| White | 2,110<br>(78%) | 659 (75%) | 127 (84%) | 34,266<br>(79%) |  |
| Black or<br>African<br>American | 393 (14%) | 141 (16%) | 17 (11%) | 5,415<br>(13%) |  |
| American<br>Indian or<br>Alaska Native | 20 (0.7%) | 7 (0.8%) | 1 (0.7%) | 258<br>(0.6%) |  |
| Native<br>Hawaiian or<br>Pacific<br>Islander | 2 (<0.1%) | 1 (0.1%) | 0 (0%) | 33<br>(<0.1%) |  |

| **Variable** | **Naproxen**<br>**n = 2,715** | **Diclofena**<br>**c**<br>**n = 879** | **Etodolac**<br>**n = 151** | **No**<br>**NSAID**<br>**n =**<br>**43,246** | **p-value** |
| --- | --- | --- | --- | --- | --- |
| Asian | 39 (1.4%) | 14 (1.6%) | 0 (0%) | 1,210<br>(2.8%) |  |
| Multiracial | 118 (4.3%) | 43 (4.9%) | 5 (3.3%) | 1,291<br>(3.0%) |  |
| Unknown | 33 (1.2%) | 14 (1.6%) | 1 (0.7%) | 773<br>(1.8%) |  |
| **Education**<br>**(years)** | 16 (13-18) | 16 (13-18) | 16 (12-18) | 16 (12-18) | 0.4 |
| **MoCA score**<br>**(Education-**<br>**corrected)** | 25 (20-27) | 25 (20-27) | 25 (21-28) | 24 (18-27) | **<0.001** |
| **Cognitive**<br>**Status**<br>**(demented)** | 889 (33%) | 252 (29%) | 54 (36%) | 18,815<br>(44%) | **<0.001** |
| **Evidence of Alzheimer's disease** |  |  |  |  | **<0.001** |
| Normal cognition | 467 (17%) | 164 (19%) | 25 (17%) | 9,014<br>(21%) |  |
| Diagnosis (presumptive) | 1,013<br>(37%) | 313 (36%) | 63 (42%) | 19,203<br>(44%) |  |
| No Diagnosis | 1,235<br>(45%) | 402 (46%) | 63 (42%) | 15,029<br>(35%) |  |
| **Elevated Tau in Cerebrospinal Fluid** |  |  |  |  | 0.3 |
| Positive | 36 (40%) | 13 (45%) | 2 (100%) | 615 (47%) |  |
| Negative | 53 (60%) | 16 (55%) | 0 (0%) | 691 (53%) |  |
| **Low Amyloid Cerebrospinal Fluid** |  |  |  |  | 0.2 |
| Positive | 41 (44%) | 13 (45%) | 2 (100%) | 716 (52%) |  |
| Negative | 53 (56%) | 16 (55%) | 0 (0%) | 674 (48%) |  |
| **Cerebrospinal Fluid- Status** |  |  |  |  | 0.6 |
| Elevated Tau<br>and low<br>Amyloid | 27 (31%) | 9 (33%) | 2 (100%) | 504 (40%) |  |
| Normal Tau<br>and normal<br>Amyloid | 41 (47%) | 12 (44%) | 0 (0%) | 525 (42%) |  |
| Only low<br>Amyloid | 11 (13%) | 3 (11%) | 0 (0%) | 135 (11%) |  |
| Only<br>elevated Tau | 8 (9.2%) | 3 (11%) | 0 (0%) | 87 (7.0%) |  |

After matching, baseline characteristics were well balanced between NSAID users and their matched non-user controls (***Supplementary Tables 2–4***). As shown in ***Figure 1***, both diclofenac and naproxen use were associated with significantly lower prevalence of dementia and AD compared with matched non-users (diclofenac: dementia 28.6% vs 35.0%, *P* value <.05, AD 43.3% vs. 51.1% *P* value < .01; naproxen: dementia 31.2% vs 38.6%, *P* value < .001; AD 44.8% vs 51.8%, *P* value < .001). The matched etodolac group was the smallest, including 125 participants in total, with 46 in the dementia subgroup and 62 in the AD subgroup (***Supplementary Table 4***). Although the prevalence of dementia (36.8% vs 41.6%) and AD (49.6% vs 56.0%) was lower among etodolac users than among matched non-users, these differences did not reach statistical significance (P values = .52 and .38, respectively).

**Figure 1.**
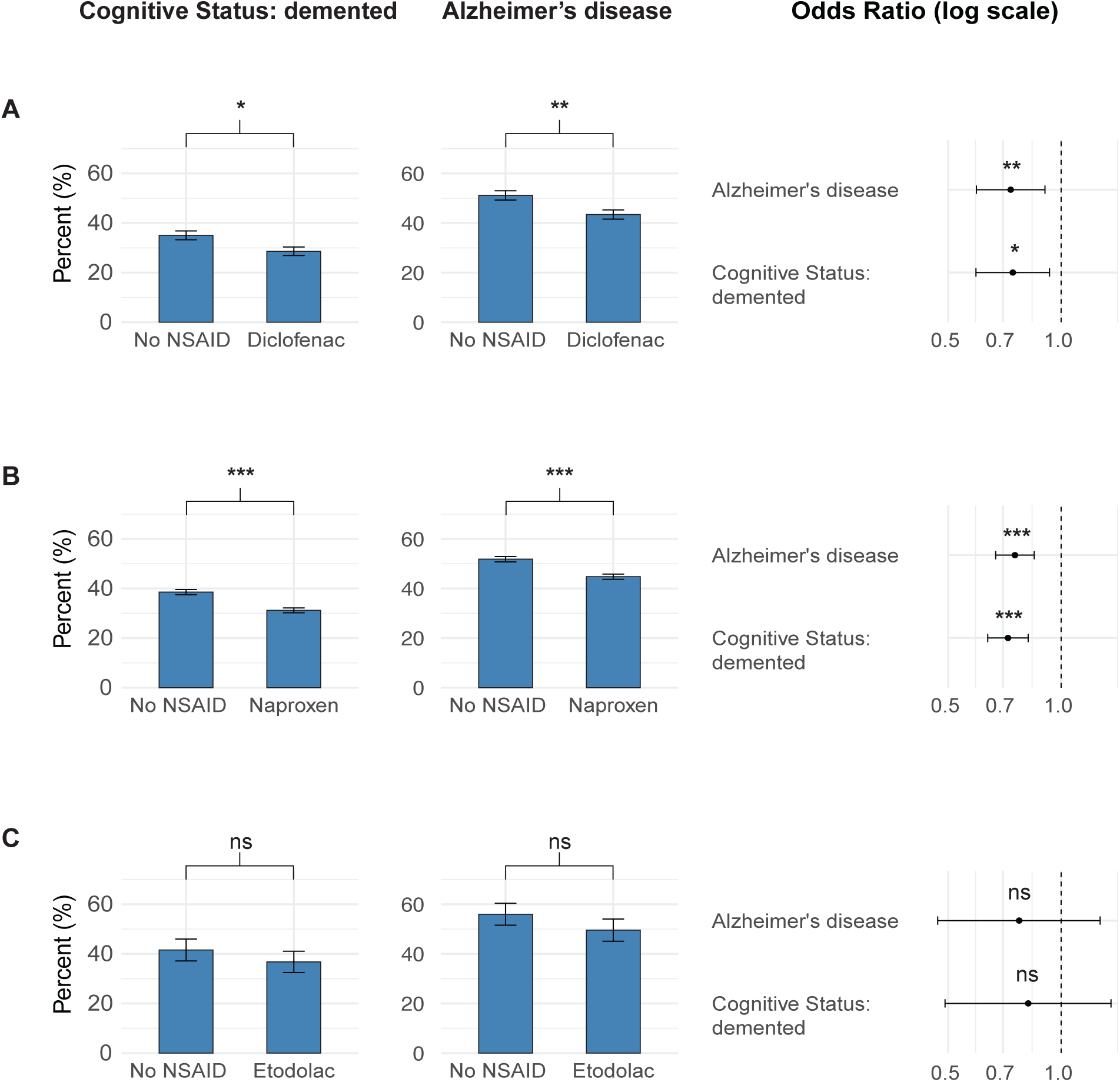
Prevalence of dementia or Alzheimer’s disease among NSAID users compared to participants with no reported NSAID use. Each row compares the use of a specific NSAID (A: Diclofenac, B: Naproxen, or C: Etodolac) with each matched cohort that does not use NSAIDs (No NSAID). The left and middle columns show corresponding prevalence rates for cognitive status (demented) and Alzheimer’s disease, respectively, by group. The right column displays forest plots of odds ratios and 95% confidence intervals for the association between NSAID use and either Alzheimer’s disease or cognitive status. Significance levels are indicated as follows: n.s.= not significant, p < 0.05 (*), p < 0.01 (**), and p < 0.001 (***).

Corresponding odds ratios were significantly lower for both diclofenac (dementia: OR = 0.74, 95% CI 0.59–0.93, *P* value = .010; AD: OR = 0.73, 95% CI 0.59–0.91, *P* value = .004) and naproxen (dementia: OR = 0.72, 95% CI 0.64–0.82, *P* value < .001; AD: OR = 0.75, 95% CI 0.67–0.85, *P* value <.001), indicating a lower likelihood of dementia and AD compared to non-users. In contrast, etodolac showed no significant association with either dementia (OR = 0.82, 95% CI 0.49–1.36, *P* value = .44) or AD (OR = 0.77, 95% CI 0.47–1.27, *P* value = .31).

### 3.3. NSAID use and cognitive performance

We further expected diclofenac users to show better cognitive performance as measured by MoCA scores (***Figure 2***). In the total cohort, as well as among participants with dementia, naproxen users demonstrated higher median MoCA scores than non-users [total cohort: 25.0 (IQR 19-27) versus 25.0 (IQR 21-28), *P* value < .01; dementia subgroup: 13.0 (IQR 7-19) versus 15.5 (IQR 9-20), *P* value < .01; AD subgroup: 19.5 (IQR 13-23) versus 18.0 (IQR 11-23), *P* value = .09]. Only the differences between the total cohort and the dementia subgroup were statistically significant, consistent with the lower dementia prevalence observed in these groups. Of note, for diclofenac non-users versus users, median MoCA scores were higher in the total cohort (24 (IQR 20-27) versus 25 (IQR 21-28), *P* value =.12), in the dementia group (13.0 (IQR 7-19) versus 15.0 (IQR 10-20), *P* value = .06) and in the AD group (18 (IQR 11-23) versus 20 (IQR 14-23), *P* value=.07). Still, these differences did not reach statistical significance.

**Figure 2.**
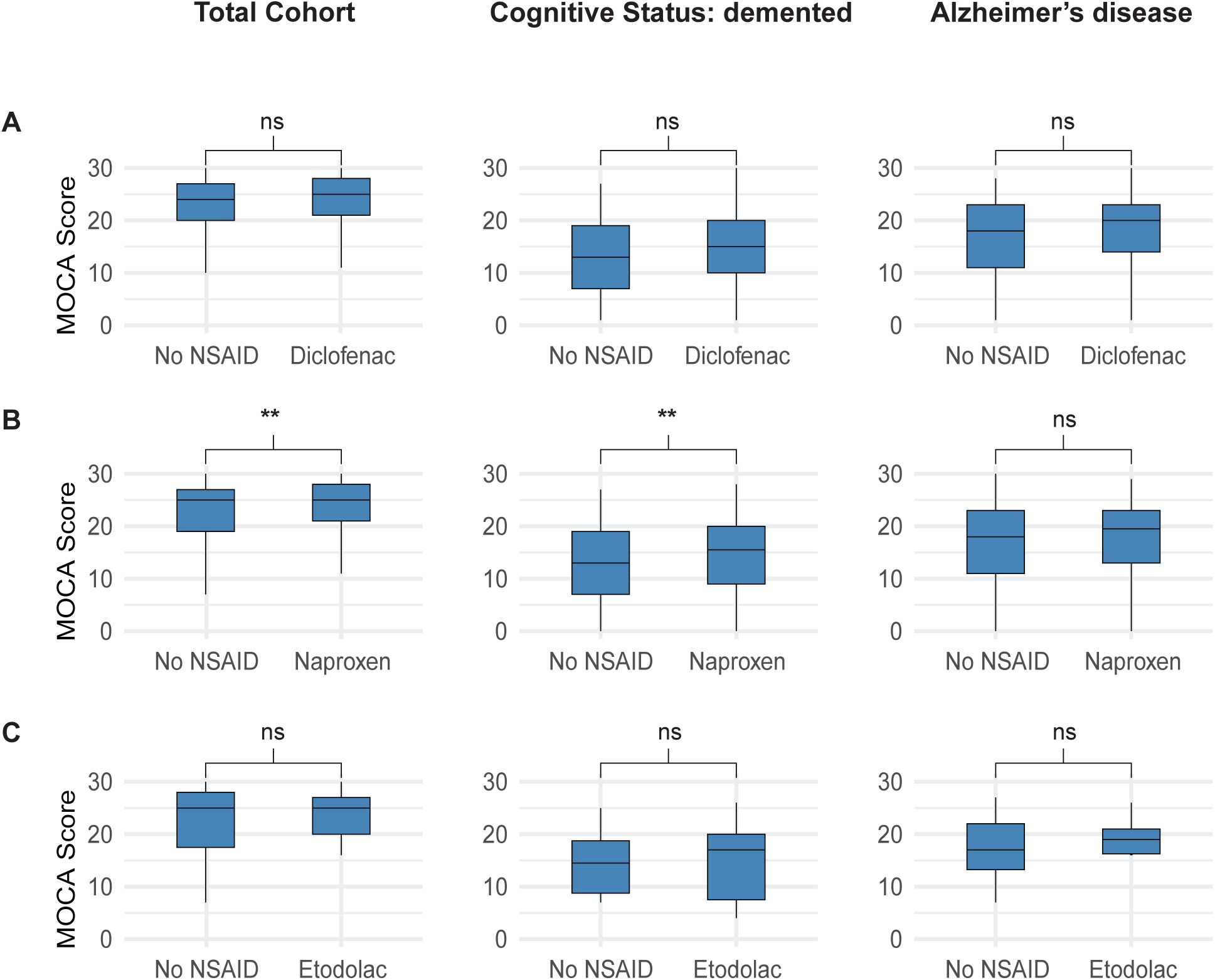
MoCA Scores by NSAID Across Diagnostic Subgroups. Each boxplot compares MoCA scores between participants using a specific NSAID (A: Diclofenac, B: Naproxen, or C: Etodolac) and matched participants not using NSAIDs (No NSAID). Rows represent the NSAID types, and columns represent diagnostic subgroups: total cohort (left), participants with dementia (middle), and participants with Alzheimer’s disease diagnosis (right). p-values from Wilcoxon rank-sum tests are shown above each comparison. Significance levels are indicated as follows: n.s. not significant, p < 0.05 (*), p < 0.01 (**), and p < 0.001 (***).

Similar results were observed for the etodolac group, where MoCA scores were slightly higher across all subgroups, while the differences did not reach statistical significance [total cohort: 25.0 (IQR 17.5-28) versus 25.0 (IQR 20-27), *P* value = .73; dementia: 17.0 (IQR 12.5) versus 14.5 (IQR 10), *P* value = .88; AD: 19.0 (IQR 16.25-21) versus 17.0 (IQR 13.25-22), *P* value = .85].

### 3.4. NSAID use and longitudinal effects on cognition

To further assess the longitudinal effects of NSAID use, we modeled MoCA score trajectories over time, hypothesizing that diclofenac use would be associated with a slower rate of cognitive decline (***Figure 3***). Across all groups, cognitive function declined over the modeled 15-year period. Interestingly, the rate of decline differed by NSAID exposure. Participants using diclofenac exhibited significantly slower cognitive decline than non-users (slope: −0.23 vs −0.38 MoCA points per year; *P* < .01). Naproxen and etodolac users showed intermediate rates of decline (−0.32 and −0.29 per year, respectively), although these did not reach statistical significance. Participants reporting the use of multiple NSAIDs demonstrated a similarly attenuated decline (−0.25 per year), which was not significantly different from diclofenac alone.

**Figure 3.**
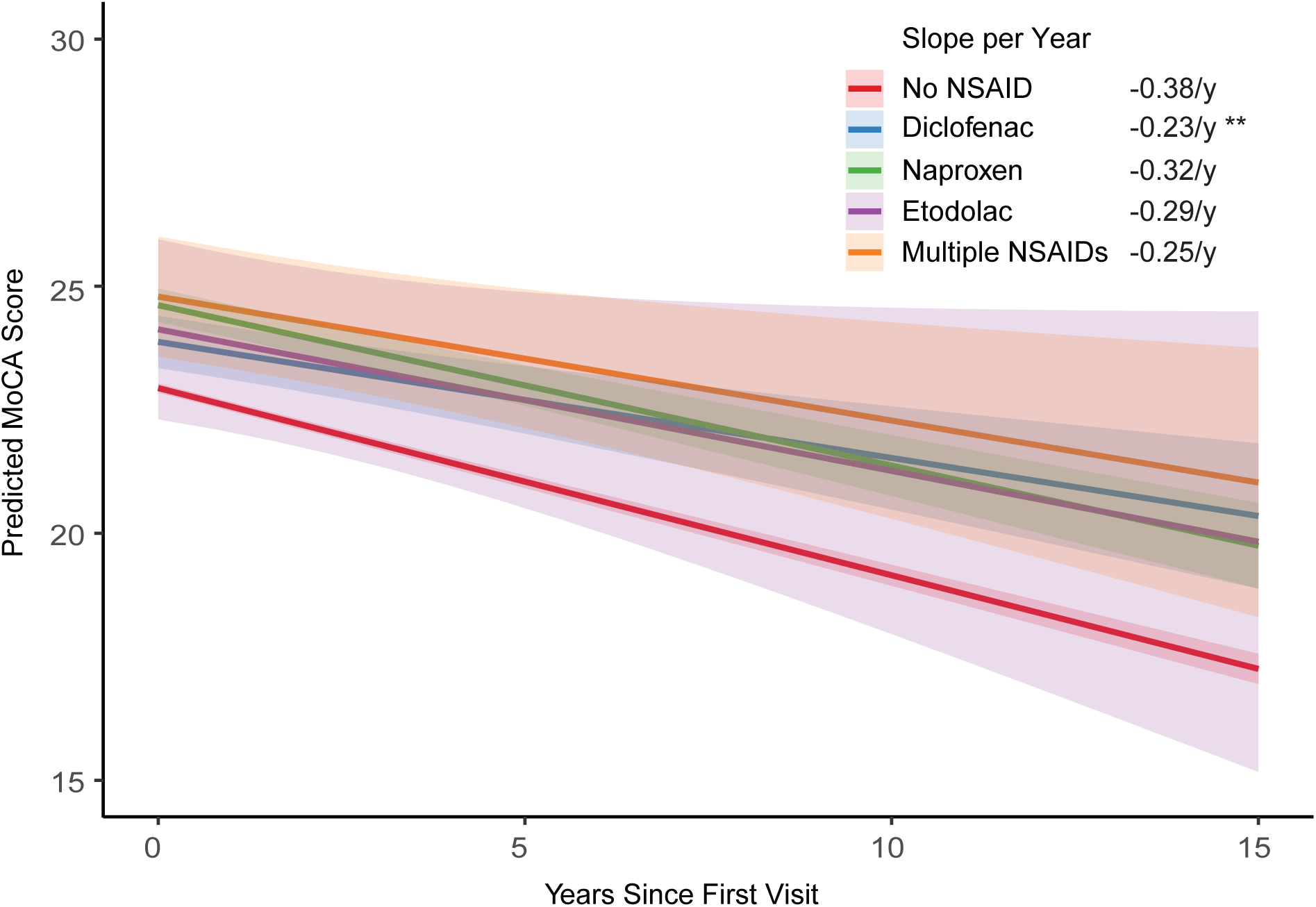
MoCA Score Trajectories Over Time by NSAID Use. The figure shows predicted longitudinal MoCA scores over 15 years of follow-up for participants grouped by NSAID use. Each colored line represents a different NSAID category: No NSAID (reference), Diclofenac, Naproxen, Etodolac, or Multiple NSAIDs. Shaded ribbons indicate 95% confidence intervals. Slopes (annual change in MoCA score) are shown in the legend with significance levels: p < 0.05 (*), p < 0.01 (**), and p < 0.001 (***).

### 3.5. Diagnostic criteria and UDS versions

To account for differing diagnostic criteria, we repeated the analysis in UDS 3.0–3.2 participants (n = 24,284; ***Supplementary Table 5***). Only naproxen replicated the association with lower dementia and AD prevalence, whereas diclofenac and etodolac showed similar but non-significant trends, likely due to smaller sample sizes and diagnostic variability.

## 4. Discussion

Across a large NACC cohort, we found evidence that NSAID exposure was associated with a lower prevalence of dementia and AD, with compound-specific effects. Most notably, diclofenac use was associated with a slower rate of cognitive decline in a longitudinal model. A cross-sectional association between NSAID intake and lower prevalence of dementia and AD was also observed for naproxen, while etodolac showed similar but non-significant directional trends.

### Epidemiological evidence

Epidemiological studies have repeatedly hinted at reduced AD risk among NSAID users, although results vary widely by compound and study design [30]. Meta-analytic evidence supports a modest overall risk reduction, while subgroup analyses often fail to identify consistent benefits for individual drugs [20]. Similarly, large case–control analyses have suggested protective associations, most prominently for ibuprofen, yet effects across NSAID classes remain inconsistent [31]. By contrast, randomized controlled trials have largely reported negative results. A Cochrane review found no cognitive benefit for various NSAIDs or COX-2 inhibitors and noted increased adverse events [32]. Our longitudinal model analysis differs from these studies in two important ways: it predicts cognitive trajectories over a much longer time span (15 years), and it includes individuals across a broader spectrum of disease stages, including early symptomatic and asymptomatic patients. This extended observational window may have revealed associations missed in shorter studies of more advanced, symptomatic patients. These differences underscore the importance of timing, compound specificity, and exposure duration when evaluating NSAIDs as potential disease-modifying agents in AD.

Our findings confirm prior modeling work by Rivers-Auty et al. (2020) based on data from the Alzheimer’s Disease Neuroimaging Initiative, demonstrating long-term diclofenac exposure was associated with reduced dementia risk over time [33]. Similar to our study, this analysis emphasized temporal dynamics rather than cross-sectional comparisons, suggesting that sustained modulation of inflammatory pathways may be required to detect protective effects [33]. The convergence of results across independent cohorts strengthens the evidence that diclofenac, unlike many other NSAIDs, may exert meaningful long-term effects on neurodegenerative outcomes.

### Mechanisms

The compound-specific patterns observed in our study raise important questions about underlying mechanisms. Diclofenac, a phenylacetic acid derivative of the NSAID class with non-selective COX-1/2 inhibition, has been proposed to exert neuroprotective effects through several pathways [6]. Studies suggest that diclofenac can cross the BBB and modulate microglial activation states, including partial inhibition of the NLRP3 inflammasome, and regulation of nuclear factor kappa-light-chain-enhancer of activated B cells (NF-κB) and tumor necrosis factor alpha (TNF-α) signaling pathways [6]. Microglia play a central role in neurodegeneration, transitioning from homeostatic states to disease-associated microglia characterized by heightened inflammatory signaling, altered phagocytic capacity, and interactions with Aβ and tau pathology [34]. While acute microglial activation may initially be protective, chronic or dysregulated activation is thought to contribute to synaptic dysfunction and neuronal loss [35]. In this context, long-term attenuation of maladaptive microglial inflammasome activity, rather than complete immune suppression, may represent a disease-modifying strategy. Together, these pathways converge on innate immune regulation and neuroinflammatory signaling, which are increasingly implicated in the pathophysiology of AD and could provide a plausible mechanistic basis for the longitudinal effects observed in our study.

The significant cross-sectional associations observed for naproxen were unexpected, and the results did not show robust longitudinal effects on cognitive decline. Naproxen is a propionic acid derivative NSAID that primarily acts as a traditional non-selective COX-1/2 inhibitor, reducing prostaglandin E2 synthesis and downstream cytokine signaling. Its penetration into the CNS is limited, and evidence for modulation of neuroinflammatory or neurodegenerative processes is inconsistent [36,37]. Nevertheless, naproxen is frequently prescribed for chronic inflammatory conditions such as rheumatoid arthritis, which are themselves associated with elevated systemic inflammatory burden and increased dementia risk [38]. More recent studies have implicated the peripheral immune system in AD pathophysiology, indicating that peripheral immune cells infiltrate the CNS, thus contributing to maladaptive and chronic microglial dysfunction [12–15]. Prolonged suppression of peripheral inflammation may therefore indirectly influence central neuroinflammatory processes.

Etodolac, an indole acetic acid derivative NSAID with preferential COX-2 inhibition, exhibited directionally similar but non-significant associations in our analyses [39,40]. Pharmacologically, etodolac exhibits limited CNS penetration and a narrower anti-inflammatory profile than diclofenac. However, emerging metabolomic data indicate that etodolac can modulate inflammatory lipid pathways implicated in neuroinflammatory signaling [39]. These observations raise the possibility that etodolac may influence CNS inflammation indirectly via peripheral metabolic networks. Accordingly, the lack of significant associations in our cohort is likely attributable to the small sample size and should be interpreted with caution and not as definitive evidence of biological inactivity.

The relationship between inflammation and neurodegeneration is complex and likely involves dynamic interactions between central and peripheral immune systems. Multiple sclerosis (MS) provides a conceptual parallel, with inflammation and neurodegeneration coexisting throughout the disease course rather than representing strictly sequential phases [41–43]. In relapsing–remitting disease, inflammatory relapses are characterized by BBB disruption and immune cell infiltration, leading to subsequent neuroinflammation and demyelination. In progressive stages, neuronal loss and brain atrophy co-exist with persistent compartmentalized inflammation in the form of chronic active lesions, characterized by an inactive gliotic core surrounded by a rim of activated microglia and macrophages [41–43]. While effective therapies exist for relapsing disease, options for progressive stages remain limited and are most effective in patients with ongoing inflammatory activity [44]. This underscores that inflammation and neurodegeneration are interconnected and dynamic processes that evolve in MS [45]. In AD, similar interactions between systemic inflammation, microglial activation, and pathological protein deposition may shape disease trajectories. NSAID-sensitive pathways, particularly those involving inflammasome signaling, therefore represent targets at the intersection of inflammatory and neurodegenerative mechanisms.

### Caveats of Cross-Sectional Cognitive Performance

Only naproxen users had significantly higher cross-sectional MoCA scores, whereas diclofenac and etodolac users showed small, non-significant increases compared with no NSAID users. These findings likely reflect limited power, particularly for etodolac, and variability in MoCA scores. We acknowledge the limited interpretability of cross-sectional comparisons of the MoCA. It may be more informative as a longitudinal measure of cognitive trajectory rather than as a one-time diagnostic marker [46]. Still, the tendency toward higher MoCA scores among NSAID users is consistent with the idea that modulation of neuroinflammatory pathways may help preserve global cognitive function.

### Limitations

Several limitations should be considered. First, the retrospective design precludes causal inference and may introduce selection bias. Second, diagnostic precision was limited: dementia and AD classifications in the NACC dataset are predominantly clinical and often lack biomarker confirmation, resulting in heterogeneous diagnostic groups. CSF data were available only for a subset, restricting assessment of Aβ and tau. Consequently, we could not robustly assess associations between NSAID exposure and Aβ or tau, an important limitation given that anti-inflammatory effects may vary depending on underlying pathology.

Third, NSAID exposure was assessed using medication inventory data, with no reliable information on duration, dosage, frequency, or adherence, thereby precluding assessment of dose–response relationships. Fourth, our finding that diclofenac no longer showed a statistically significant association with lower dementia prevalence when the analysis was restricted to the most recent UDS versions (3.0 and 3.2) suggests that our results may not be robust across different diagnostic frameworks. Finally, although our findings suggest that specific NSAID-related pathways may be biologically relevant to neurodegeneration, chronic NSAID use is not clinically feasible as a long-term preventive strategy due to well-established adverse effects, including renal, gastrointestinal, and cardiovascular toxicity [17]. Accordingly, the relevance of these findings lies less in direct therapeutic translation and more in the identification of downstream inflammatory pathways that may enable safer, more targeted interventions.

### Outlook and Future Directions

Our findings suggest that specific anti-inflammatory mechanisms, particularly those activated by diclofenac, may influence cognitive decline. Clarifying the pharmacological differences among NSAIDs could inform the development of more targeted neuroprotective strategies. Approaches such as selective NLRP3 inflammasome inhibitors or microglia-modulating substances may offer central anti-inflammatory benefits while avoiding the systemic risks of chronic NSAID use.

Future studies should prioritize biomarker-defined AD cohorts with confirmed Aβ and tau pathology to increase diagnostic precision and treatment effects. Inflammatory biomarkers may further help identify subgroups most likely to benefit.

Revisiting prior NSAID trials using modern biomarker stratification or by focusing on preclinical or pathology-positive populations may reveal treatment effects that were previously masked by diagnostic heterogeneity and guide future preventive or disease-modifying clinical studies.

## Data Availability

All data produced in the present study are available upon reasonable request to the authors.

## Acknowledgements/Conflicts/Funding Sources/Consent Statement

Acknowledgments

The Alzheimer’s Disease Genetics Consortium (ADGC) is funded by a grant from the National Institute on Aging (PI, Gerard D. Schellenberg; UO1AG032984).

The NACC database is funded by NIA/NIH Grant U24 AG072122. NACC data are contributed by the NIA-funded ADRCs: P30 AG062429 (PI James Brewer, MD, PhD), P30 AG066468 (PI Oscar Lopez, MD), P30 AG062421 (PI Bradley Hyman, MD, PhD), P30 AG066509 (PI Thomas Grabowski, MD), P30 AG066514 (PI Mary Sano, PhD), P30 AG066530 (PI Helena Chui, MD), P30 AG066507 (PI Marilyn Albert, PhD), P30 AG066444 (PI David Holtzman, MD), P30 AG066518 (PI Lisa Silbert, MD, MCR), P30 AG066512 (PI Thomas Wisniewski, MD), P30 AG066462 (PI Scott Small, MD), P30 AG072979 (PI David Wolk, MD), P30 AG072972 (PI Charles DeCarli, MD), P30 AG072976 (PI Andrew Saykin, PsyD), P30 AG072975 (PI Julie A. Schneider, MD, MS), P30 AG072978 (PI Ann McKee, MD), P30 AG072977 (PI Robert Vassar, PhD), P30 AG066519 (PI Frank LaFerla, PhD), P30 AG062677 (PI Ronald Petersen, MD, PhD), P30 AG079280 (PI Jessica Langbaum, PhD), P30 AG062422 (PI Gil Rabinovici, MD), P30 AG066511 (PI Allan Levey, MD, PhD), P30 AG072946 (PI Linda Van Eldik, PhD), P30 AG062715 (PI Sanjay Asthana, MD, FRCP), P30 AG072973 (PI Russell Swerdlow, MD), P30 AG066506 (PI Glenn Smith, PhD, ABPP), P30 AG066508 (PI Stephen Strittmatter, MD, PhD), P30 AG066515 (PI Victor Henderson, MD, MS), P30 AG072947 (PI Suzanne Craft, PhD), P30 AG072931 (PI Henry Paulson, MD, PhD), P30 AG066546 (PI Sudha Seshadri, MD), P30 AG086401 (PI Erik Roberson, MD, PhD), P30 AG086404 (PI Gary Rosenberg, MD), P20 AG068082 (PI Angela Jefferson, PhD), P30 AG072958 (PI Heather Whitson, MD), P30 AG072959 (PI James Leverenz, MD).

## Consent Statement

This study did not involve direct human or animal subjects. All data were obtained from NACC, where informed consent had been obtained; therefore, no additional ethical approval or consent was required for this study.

## References

1. Wolters FJ, Ikram MA. Epidemiology of Dementia: The Burden on Society, the Challenges for Research. Biomark Alzheimers Dis Drug Dev. 2021;1750:3–14.

2. Nichols E, Steinmetz JD, Vollset SE, Fukutaki K, Chalek J, Abd-Allah F, et al. Estimation of the global prevalence of dementia in 2019 and forecasted prevalence in 2050: an analysis for the Global Burden of Disease Study 2019. Lancet Public Health. 2022;7(2):e105–e125. doi:10.1016/S2468-2667(21)00249-8

3. Reuben DB, Kremen S, Maust DT. Dementia Prevention and Treatment: A Narrative Review. JAMA Intern Med. 2024;184(5):563. doi:10.1001/jamainternmed.2023.8522

4. Braak H, Braak E. Neuropathological stageing of Alzheimer-related changes. Acta Neuropathol (Berl). 1991;82(4):239–259. doi:10.1007/BF00308809

5. Stopschinski BE, Diamond MI. The prion model for progression and diversity of neurodegenerative diseases. Lancet Neurol. 2017;16(4):323–332. doi:10.1016/s1474-4422(17)30037-6

6. Stopschinski BE, Weideman RA, McMahan D, Jacob DA, Little BB, Chiang HS, et al. Microglia as a cellular target of diclofenac therapy in Alzheimer’s disease. Ther Adv Neurol Disord. 2023;16:17562864231156674. doi:10.1177/17562864231156674

7. Jiang S, Maphis NM, Binder J, Chisholm D, Weston L, Duran W, et al. Proteopathic tau primes and activates interleukin-1â via myeloid-cell-specific MyD88- and NLRP3-ASC-inflammasome pathway. Cell Rep. 2021;36(12):109720. doi:10.1016/j.celrep.2021.109720

8. Luciunaite A, McManus RM, Jankunec M. Soluble Abeta oligomers and protofibrils induce NLRP3 inflammasome activation in microglia. J Neurochem. 2020;155:650–661.

9. Stancu IC, Lodder C, Botella Lucena P, Vanherle S, Ravé M, Terwel D, et al. The NLRP3 inflammasome modulates tau pathology and neurodegeneration in a tauopathy model. Glia. 2022;70(6):1117–1132. doi:10.1002/glia.24160

10. Ising C, Venegas C, Zhang S, Scheiblich H, Schmidt SV, Vieira-Saecker A, et al. NLRP3 inflammasome activation drives tau pathology. Nature. 2019;575(7784):669–673. doi:10.1038/s41586-019-1769-z

11. Sheng JG, Zhu SG, Jones RA. Interleukin-1 promotes expression and phosphorylation of neurofilament and tau proteins in vivo. Exp Neurol. 2000;163:388–391.

12. Jorfi M, Maaser-Hecker A, Tanzi RE. The neuroimmune axis of Alzheimer’s disease. Genome Med. 2023;15(1):6. doi:10.1186/s13073-023-01155-w

13. Chen X, Holtzman DM. Emerging roles of innate and adaptive immunity in Alzheimer’s disease. Immunity. 2022;55(12):2236–2254. doi:10.1016/j.immuni.2022.10.016

14. Rossi B, Santos-Lima B, Terrabuio E, Zenaro E, Constantin G. Common Peripheral Immunity Mechanisms in Multiple Sclerosis and Alzheimer’s Disease. Front Immunol. 2021;12. doi:10.3389/fimmu.2021.639369

15. Johnson AM, Lukens JR. Emerging roles for innate and adaptive immunity in tauopathies. Cell Rep. 2025;44(9):116232. doi:10.1016/j.celrep.2025.116232

16. Gasparini L, Ongini E, Wenk G. Non-steroidal Anti-Inflammatory Drugs (NSAIDs) in Alzheimer’s Disease: Old and New Mechanisms of Action. J Neurochem. 2004;91(3):521–536.

17. Ghlichloo I, Gerriets V. Nonsteroidal Anti-Inflammatory Drugs (NSAIDs. Published online 2022.

18. Stuve O, Weideman RA, McMahan DM. Diclofenac reduces the risk of Alzheimer’s disease: a pilot analysis of NSAIDs in two US veteran populations. Ther Adv Neurol Disord. 2020;13:1756286420935676.

19. Landi F, Cesari M, Onder G. Non-steroidal anti-inflammatory drug (NSAID) use and Alzheimer disease in community-dwelling elderly patients. Am J Geriatr Psychiatry. 2003;11:179–185.

20. Zhang C, Wang Y, Wang D. NSAID exposure and risk of Alzheimer’s disease: an updated meta-analysis from cohort studies. Front Aging Neurosci. 2018;10:83.

21. Vlad SC, Miller DR, Kowall NW, Felson DT. Protective effects of NSAIDs on the development of Alzheimer disease. Neurology. 2008;70(19):1672–1677. doi:10.1212/01.wnl.0000311269.57716.63

22. Group AR, Lyketsos CG, Breitner JC. Naproxen and celecoxib do not prevent AD in early results from a randomized controlled trial. Neurology. 2007;68:1800–1808.

23. Reines SA, Block GA, Morris JC. Rofecoxib: no effect on Alzheimer’s disease in a 1-year, randomized, blinded, controlled study. Neurology. 2004;62:66–71.

24. Aisen PS, Schafer KA, Grundman M, Pfeiffer E, Sano M, Davis KL, et al. Effects of Rofecoxib or Naproxen vs Placebo on Alzheimer Disease Progression: A Randomized Controlled Trial. JAMA. 2003;289(21):2819–2826. doi:10.1001/jama.289.21.2819

25. Jack CR, Andrews JS, Beach TG, Buracchio T, Dunn B, Graf A, et al. Revised criteria for diagnosis and staging of Alzheimer’s disease: Alzheimer’s Association Workgroup. Alzheimers Dement. 2024;20(8):5143–5169. doi:10.1002/alz.13859

26. American Psychiatric Association. Diagnostic and Statistical Manual of Mental Disorders, 4th Ed. American Psychiatric Publishing, Inc.; 1994:xxvii, 886.

27. McKhann G, Drachman D, Folstein M, Katzman R, Price D, Stadlan EM. Clinical diagnosis of Alzheimer’s disease: Report of the NINCDS-ADRDA Work Group* under the auspices of Department of Health and Human Services Task Force on Alzheimer’s Disease. Neurology. 1984;34(7):939–939. doi:10.1212/WNL.34.7.939

28. Sperling RA, Aisen PS, Beckett LA, Bennett DA, Craft S, Fagan AM, et al. Toward defining the preclinical stages of Alzheimer’s disease: Recommendations from the National Institute on Aging-Alzheimer’s Association workgroups on diagnostic guidelines for Alzheimer’s disease. Alzheimers Dement. 2011;7(3):280–292. doi:10.1016/j.jalz.2011.03.003

29. Austin PC, Yu AYX, Vyas MV, Kapral MK. Applying Propensity Score Methods in Clinical Research in Neurology. Neurology. 2021;97(18):856–863. doi:10.1212/WNL.0000000000012777

30. Gasparini L, Ongini E, Wenk G. Non-steroidal Anti-Inflammatory Drugs (NSAIDs) in Alzheimer’s Disease: Old and New Mechanisms of Action. J Neurochem. 2004;91(3):521–536.

31. Vlad SC, Miller DR, Kowall NW. Protective effects of NSAIDs on the development of Alzheimer disease. Neurology. 2008;70:1672–1677.

32. Jaturapatporn D, Isaac MG, McCleery J. Aspirin, steroidal and non-steroidal anti-inflammatory drugs for the treatment of Alzheimer’s disease. Cochrane Database Syst Rev. 2012;2:006378.

33. Rivers-Auty J, Mather AE, Peters R, Lawrence CB, Brough D, Alzheimer’s Disease Neuroimaging Initiative. Anti-inflammatories in Alzheimer’s disease—potential therapy or spurious correlate? Brain Commun. 2020;2(2):fcaa109. doi:10.1093/braincomms/fcaa109

34. Krasemann S, Madore C, Cialic R, Baufeld C, Calcagno N, El Fatimy R, et al. The TREM2-APOE Pathway Drives the Transcriptional Phenotype of Dysfunctional Microglia in Neurodegenerative Diseases. Immunity. 2017;47(3):566–581.e9. doi:10.1016/j.immuni.2017.08.008

35. Yang G, Xu X, Gao W, Wang X, Zhao Y, Xu Y. Microglia-orchestrated neuroinflammation and synaptic remodeling: roles of pro-inflammatory cytokines and receptors in neurodegeneration. Front Cell Neurosci. 2025;19:1700692. doi:10.3389/fncel.2025.1700692

36. Kim S, Chang WE, Kumar R, Klimov DK. Naproxen Interferes with the Assembly of Aâ Oligomers Implicated in Alzheimer’s Disease. Biophys J. 2011;100(8):2024–2032. doi:10.1016/j.bpj.2011.02.044

37. Meyer PF, Tremblay-Mercier J, Leoutsakos J. INTREPAD: a randomized trial of naproxen to slow progress of presymptomatic Alzheimer disease. Neurology. 2019;92:2070–2080.

38. Myasoedova E, Sattui SE, Lee J, O’Brien JT, Makris UE. Cognitive impairment in individuals with rheumatic diseases: the role of systemic inflammation, immunomodulatory medications, and comorbidities. Lancet Rheumatol. 2024;6(12):e871–e880. doi:10.1016/S2665-9913(24)00190-5

39. Sebaa R, AlMalki RH, Sukkarieh H, Dahabiyeh LA, Mogren MA, Arafat T, et al. Systemic Metabolic Alterations Induced by Etodolac in Healthy Individuals. Pharmaceuticals. 2025;18(8):1155. doi:10.3390/ph18081155

40. Tachibana M, Inoue N, Yoshida E, Matsui M, Ukai Y, Yano J. Anti-Inflammatory Effect and Low Ulcerogenic Activity of Etodolac, a Cyclooxygenase-2 Selective Non-Steroidal Anti-Inflammatory Drug, on Adjuvant-Induced Arthritis in Rats. Pharmacology. 2003;68(2):96–104. doi:10.1159/000069536

41. Milo R, Korczyn AD, Manouchehri N, Stüve O. The temporal and causal relationship between inflammation and neurodegeneration in multiple sclerosis. Mult Scler J. 2020;26(8):876–886. doi:10.1177/1352458519886943

42. McGinley MP, Goldschmidt CH, Rae-Grant AD. Diagnosis and Treatment of Multiple Sclerosis: A Review. JAMA. 2021;325(8):765. doi:10.1001/jama.2020.26858

43. Boutitah-Benyaich I, Eixarch H, Villacieros-Álvarez J, Hervera A, Cobo-Calvo Á, Montalban X, et al. Multiple sclerosis: molecular pathogenesis and therapeutic intervention. Signal Transduct Target Ther. 2025;10(1):324. doi:10.1038/s41392-025-02415-4

44. Chataway J, Williams T, Li V, Marrie RA, Ontaneda D, Fox RJ. Clinical trials for progressive multiple sclerosis: progress, new lessons learned, and remaining challenges. Lancet Neurol. 2024;23(3):277–301. doi:10.1016/S1474-4422(24)00027-9

45. Hoehne C, Stuve O, Stopschinski BE. Tau in Multiple Sclerosis: A Review of Therapeutic Potential. Curr Treat Options Neurol. 2025;27(1):14. doi:10.1007/s11940-024-00820-8

46. Bernier PJ, Gourdeau C, Carmichael P, Beauchemin J, Voyer P, Hudon C, et al. It’s all about cognitive trajectory: Accuracy of the cognitive charts– MOCA in normal aging, MCI, and dementia. J Am Geriatr Soc. 2023;71(1):214–220. doi:10.1111/jgs.18029

