## Supplement for "NSAID use is associated with lower dementia and Alzheimer’s disease prevalence and slower cognitive decline: A retrospective longitudinal analysis of the NACC cohort"

ALZHEIMER'S & DEMENTIA

Carolin Hoehne, M.D. [1]

Victor Salinas, M.D., Ph.D.[2]

Afsaneh, Shirani, M.D., MSCI [3,4]

Olaf Stuve, M.D., Ph.D.[5,6,7]

Barbara Elena Stopschinski, M.D. [5,6,8]

Address

[1] Department of Neurology, Charité Universitätsmedizin Berlin, Bonhoefferweg 3, 10117 Berlin, Germany.

[2] Rocky Mountain Movement Disorders Center, Englewood, CO, USA.

[3] Saint Luke's Marion Bloch Neuroscience Institute, Kansas City, MO, USA.

[4] Department of Neurology, University of Missouri-Kansas City, Kansas City, MO, USA.

[5] Department of Neurology, University of Texas Southwestern Medical Center, Dallas, TX, USA.

[6] Peter O'Donnell Brain Institute, University of Texas Southwestern Medical Center, Dallas, TX, USA.

[7] Neurology Section, Dallas VA Medical Center, 4500 South Lancaster Road, Dallas, TX, USA.

[8] Center for Alzheimer's and Neurodegenerative Diseases, University of Texas Southwestern Medical Center, Dallas, TX, USA.

Correspondence

<u>**List of Content**</u>

**sFigure 1 Form D1 (Clinical Questionnaire) of the USD Version3.0. 9**

**sTable 1 List of NACC variables included in the analysis.**

| NACC-Variable | Description | NACC Codes |
|---|---|---|
| **NACC Visit data** | | |
| NACCID | Participant Study ID | Unique value |
| NACCVNUM | Visit Number | Longitudinal Counting |
| NACCFDYS | Days from initial visit to most recent | Longitudinal Counting |
| FORMVER | Form version number | V1.2, V2, V3.0, V3.2 |
| **Demographics** | | |
| NACCAGE | Participants Age at Visit | Longitudinal Counting |
| SEX | Participant’s Sex | 1 = Male 2 = Female |
| NACCNIHR | Derived NIH race definition | 1 = White<br>2 = Black or African American<br>3 = American Indian or Alaska Native<br>4 = Native Hawaiian or Pacific Islander<br>5 = Asian<br>6 = Multiracial<br>99 = Unknown or ambiguous |
| EDUC | Years of education | 0 – 36<br>99 = Unknown |

| NACC-Variable | Description | NACC Codes |
|---|---|---|
| **Clinical status/ Outcomes** | | |
| NACCALZD | Presumptive etiologic diagnosis of the cognitive disorder — Alzheimer's disease | 0 = No (assumed assessed and found not present)<br>1 = Yes<br>8 = No cognitive impairment |
| DEMENTED | Met criteria for dementia | 0 = No<br>1 = Yes |
| NACCMOCA | MoCA Total Score — corrected for education | 0 – 30<br>88 = Item(s) or whole test not administered<br>99= Years of education missing/unknown<br>-4 = Not available: UDS form submitted did not collect data in this way, or a skip pattern precludes response to this question |
| CSFTAU | Abnormally elevated Tau or phospho- Tau in CSF | 0 = No<br>1 = Yes<br>8 = Unknown/ not assessed<br>-4 = Not applicable: UDS form submitted did not collect data in this way, or a skip pattern precludes response to this question |
| AMYLCSF | Abnormally low amyloid in CSF | 0 = No<br>1 = Yes<br>8 = Unknown/not assessed<br>-4 = Not applicable: UDS form submitted did not collect data in this way, or a skip pattern precludes response to this question |
| **Comorbidities** | | |

| NACC-Variable | Description | NACC Codes |
| --- | --- | --- |
| NACCTBI | History of traumatic brain injury | 0 = No<br>1 = Yes<br>9 = Unknown<br>- 4 = Not available: UDS form submitted did not collect data in this way, or a skip pattern precludes response to this question |
| HXHYPER | History or presence of hypertension | 0 = Absent<br>1 = Present<br>- 4 = Not available: UDS form submitted did not collect data in this way, or a skip pattern precludes response to this question |
| HXSTROKE | History of stroke | 0 = Absent<br>2 = Present<br>- 4 = Not available: UDS form submitted did not collect data in this way, or a skip pattern precludes response to this question |
| DEP | Depression or dysphoria in the last | 0 = No<br>1 = Yes<br>9 = Unknown<br>-4 = Not available: UDS form submitted did not collect data in this way, or a skip pattern precludes response to this question |
| BIPOLDX | Presumptive etiologic diagnosis — bipolar disorder | 0 = No (assumed assessed and found not present)<br>1 = Yes<br>-4 = Not applicable: UDS form submitted did not collect data in this way, or a skip pattern |

| NACC-Variable | Description | NACC Codes |
| --- | --- | --- |
| | | precludes response to this question |
| SCHIZOP | Presumptive etiologic diagnosis —Schizophrenia or other psychosis | 0 = No (assumed assessed and found not present)<br>1 = Yes<br>-4 = Not applicable: UDS form submitted did not collect data in this way, or a skip pattern precludes response to this question |
| ANXIET | Presumptive etiologic diagnosis — Anxiety | 0 = No (assumed assessed and found not present)<br>1 = Yes<br>-4 = Not applicable: UDS form submitted did not collect data in this way, or a skip pattern precludes response to this question |
| PTSDDX | Presumptive etiologic diagnosis — Post-traumatic stress disorder (PTSD) | 0 = No (assumed assessed and found not present)<br>1 = Yes<br>-4 = Not applicable: UDS form submitted did not collect data in this way, or a skip pattern precludes response to this question |
| OTHPSY | Presumptive etiologic diagnosis — Other psychiatric diseases | 0 = No (assumed assessed and found not present)<br>1 = Yes |
| ALCABUSE | Current alcohol abuse | 0 = No<br>1 = Yes<br>9 = Unknown<br>8 = No diagnosis of impairment due to alcohol abuse<br>-4 = Not applicable: UDS form submitted did not collect data in |

| NACC-Variable | Description | NACC Codes |
|---|---|---|
| | | this way, or a skip pattern precludes response to this question |
| CANCER | Cancer present in the last 12 months (excluding non-melanoma skin cancer), primary or metastatic | 0 = No<br>1 = Yes, primary/ non-metastatic<br>2 = Yes, metastatic<br>8 = Not assessed<br>-4 = Not available: UDS form submitted did not collect data in this way, or a skip pattern precludes response to this question |
| DIABETES | Diabetes not | 0 = Absent<br>1 = Recent/Active<br>2 = Remote/Inactive<br>9 = Unknown<br>- 4 = Not available: UDS form submitted did not collect data in this way, or a skip pattern precludes response to this question |
| MYOINF | Myocardial infarct present within the past 12 month | 0 = No<br>1 = Yes<br>8 = Not assessed<br>-4 = Not available: UDS form submitted did not collect data in this way, or a skip pattern precludes response to this question |
| CONGHRT | Congestive heart failure present | 0 = No<br>1 = Yes<br>8 = Not assessed<br>-4 = Not available: UDS form submitted did not collect data in this way, or a skip pattern |

| NACC-Variable | Description | NACC Codes |
|---|---|---|
| | | precludes response to this question |
| AFIBRILL | Atrial fibrillation present | 0 = No<br>1 = Yes<br>8 = Not assessed<br>-4 = Not available: UDS form submitted did not collect data in this way, or a skip pattern precludes response to this question |
| HYPERT | Hypertension present | 0 = No<br>1 = Yes<br>8 = Not assessed<br>-4 = Not available: UDS form submitted did not collect data in this way, or a skip pattern precludes response to this question |
| HYPCHOL | Hypercholesterolemia present | 0 = No<br>1 = Yes<br>8 = Not assessed<br>-4 = Not available: UDS form submitted did not collect data in this way, or a skip pattern precludes response to this question |
| VB12DEF | Vitamin B12 deficiency present | 0 = No<br>1 = Yes<br>8 = Not assessed<br>-4 = Not available: UDS form submitted did not collect data in this way, or a skip pattern precludes response to this question |
| THYDIS | Thyroid disease present | 0 = No<br>1 = Yes |

| NACC-Variable | Description | NACC Codes |
|---|---|---|
| | | 8 = Not assessed<br>-4 = Not available: UDS form submitted did not collect data in this way, or a skip pattern precludes response to this question |
| ARTH | Arthritis present | 0 = No<br>1 = Yes<br>8 = Not assessed<br>-4 = Not available: UDS form submitted did not collect data in this way, or a skip pattern precludes response to this question |
| SLEEPAP | Sleep apnea present | 0 = No<br>1 = Yes<br>8 = Not assessed<br>-4 = Not available: UDS form submitted did not collect data in this way, or a skip pattern precludes response to this question |
| OTHCOND | Other medical conditions or procedures within the past 12 months not listed | 0 = No<br>1 = Yes<br>-4 = Not available: UDS form submitted did not collect data in this way, or a skip pattern precludes response to this question |

### sFigure 1 Form D1 (Clinical Questionnaire) of the USD Version 3.0.

NACC National Alzheimer's Coordinating Center

**INITIAL VISIT PACKET** NACC UNIFORM DATA SET (UDS)

#### Form D1: Clinician Diagnosis

ADC name: ______ Subject ID: ______ Form date: __ / __ / ____
Visit #: ___ Examiner's initials: ___

*INSTRUCTIONS: This form is to be completed by the clinician. For additional clarification and examples, see UDS Coding Guidebook for Initial Visit Packet, Form D1. Check only one box per question.*

This form is divided into three main sections:

Section 1 **Cognitive and behavioral status:** Normal cognition / MCI / dementia and dementia syndrome
Section 2 **Biomarkers, imaging, and genetics:** Neurodegenerative imaging and CSF biomarkers, imaging evidence for CVD, and known genetic mutations for AD and FTLD
Section 3 **Etiological diagnoses:** presumed etiological diagnoses for the cognitive disorder

**1. Diagnosis method — responses in this form are based on diagnosis by:**
☐ 1 A single clinician ☐ 2 A formal consensus panel ☐ 3 Other (e.g., two or more clinicians or other informal group)

**SECTION 1: Cognitive and behavioral status**

**2. Does the subject have normal cognition (global CDR=0 and/or neuropsychological testing within normal range) and normal behavior (i.e., the subject does not exhibit behavior sufficient to diagnose MCI or dementia due to FTLD or LBD)?**
☐ 0 No **(CONTINUE TO QUESTION 3)**
☐ 1 Yes **(SKIP TO QUESTION 6)**

ALL-CAUSE DEMENTIA

**The subject has cognitive or behavioral (neuropsychiatric) symptoms that meet all of the following criteria:**
- Interfere with ability to function as before at work or at usual activities?
- Represent a decline from previous levels of functioning?
- Are not explained by delirium or major psychiatric disorder?
- Include cognitive impairment detected and diagnosed through a combination of 1) history-taking and 2) objective cognitive assessment (bedside or neuropsychological testing)?

**AND**

**Impairment in one* or more of the following domains.**
- Impaired ability to acquire and remember new information
- Impaired reasoning and handling of complex tasks, poor judgment
- Impaired visuospatial abilities
- Impaired language functions
- Changes in personality, behavior, or comportment

** In the event of single-domain impairment (e.g., language in PPA, behavior in bvFTD, posterior cortical atrophy), the subject must not fulfill criteria for MCI.*

**3. Does the subject meet the criteria for dementia?**
☐ 0 No **(SKIP TO QUESTION 5)**
☐ 1 Yes **(CONTINUE TO QUESTION 4)**

**4. If the subject meets criteria for dementia, answer Questions 4a–4f below and then SKIP TO QUESTION 6.**

Based entirely on the history and examination (including neuropsychological testing), what is the cognitive/behavioral syndrome? **Select one or more as Present; all others will default to Absent in the NACC database.**

| Dementia syndrome | Present |
|---|---|
| 4a. Amnestic multidomain dementia syndrome | ☐ 1 |
| 4b. Posterior cortical atrophy syndrome (or primary visual presentation) | ☐ 1 |
| 4c. Primary progressive aphasia (PPA) syndrome<br>4c1. ☐ 1 Meets criteria for semantic PPA<br>☐ 2 Meets criteria for logopenic PPA<br>☐ 3 Meets criteria for nonfluent/agrammatic PPA<br>☐ 4 PPA other/not otherwise specified | ☐ 1 |
| 4d. Behavioral variant FTD (bvFTD) syndrome | ☐ 1 |
| 4e. Lewy body dementia syndrome | ☐ 1 |
| 4f. Non-amnestic multidomain dementia, not PCA, PPA, bvFTD, or DLB syndrome | ☐ 1 |

**5. If the subject does not have normal cognition or behavior and is not clinically demented, indicate the type of cognitive impairment below.**

MCI CORE CLINICAL CRITERIA
- Is the subject, the co-participant, or a clinician concerned about a change in cognition compared to the subject's previous level?
- Is there impairment in one or more cognitive domains (memory, language, executive function, attention, and visuospatial skills)?
- Is there largely preserved independence in functional abilities (no change from prior manner of functioning or uses minimal aids or assistance)?

Select one syndrome from 5a–5e as being Present (all others will default to Absent in the NACC database), and then **CONTINUE TO QUESTION 6**. If you select MCI below, it should meet the MCI core clinical criteria outlined above.

| Type | Present | Affected domains | No | Yes |
|---|---|---|---|---|
| 5a. Amnestic MCI, single domain (aMCI SD) | ☐ 1 | | | |
| 5b. Amnestic MCI, multiple domains (aMCI MD) | ☐ 1 | **CHECK YES for at least one additional domain (besides memory):** | | |
| | | 5b1. Language | ☐ 0 | ☐ 1 |
| | | 5b2. Attention | ☐ 0 | ☐ 1 |
| | | 5b3. Executive | ☐ 0 | ☐ 1 |
| | | 5b4. Visuospatial | ☐ 0 | ☐ 1 |

Select one syndrome from 5a–5e as being Present (all others will default to Absent in the NACC database), and then **CONTINUE TO QUESTION 6**. If you select MCI below, it should meet the MCI core clinical criteria outlined above.

| Type | Present | Affected domains | No | Yes |
|---|---|---|---|---|
| 5c. Non-amnestic MCI, single domain (naMCI SD) | ☐ 1 | **CHECK YES to indicate the affected domain:** | | |
| | | 5c1. Language | ☐ 0 | ☐ 1 |
| | | 5c2. Attention | ☐ 0 | ☐ 1 |
| | | 5c3. Executive | ☐ 0 | ☐ 1 |
| | | 5c4. Visuospatial | ☐ 0 | ☐ 1 |
| 5d. Non-amnestic MCI, multiple domains (naMCI MD) | ☐ 1 | **CHECK YES for at least two domains:** | | |
| | | 5d1. Language | ☐ 0 | ☐ 1 |
| | | 5d2. Attention | ☐ 0 | ☐ 1 |
| | | 5d3. Executive | ☐ 0 | ☐ 1 |
| | | 5d4. Visuospatial | ☐ 0 | ☐ 1 |
| 5e. Cognitively impaired, not MCI | ☐ 1 | | | |

**SECTION 2: Biomarkers, imaging, and genetics**

*Section 2 must be completed for all subjects.*

**6. Indicate neurodegenerative biomarker status, using local standards for positivity.**

| Biomarker findings | No | Yes | Unknown/ not assessed |
|---|---|---|---|
| 6a. Abnormally elevated amyloid on PET | ☐ 0 | ☐ 1 | ☐ 8 |
| 6b. Abnormally low amyloid in CSF | ☐ 0 | ☐ 1 | ☐ 8 |
| 6c. FDG-PET pattern of AD | ☐ 0 | ☐ 1 | ☐ 8 |
| 6d. Hippocampal atrophy | ☐ 0 | ☐ 1 | ☐ 8 |
| 6e. Tau PET evidence for AD | ☐ 0 | ☐ 1 | ☐ 8 |
| 6f. Abnormally elevated CSF tau or ptau | ☐ 0 | ☐ 1 | ☐ 8 |
| 6g. FDG-PET evidence for frontal or anterior temporal hypometabolism for FTLD | ☐ 0 | ☐ 1 | ☐ 8 |
| 6h. Tau PET evidence for FTLD | ☐ 0 | ☐ 1 | ☐ 8 |
| 6i. Structural MR evidence for frontal or anterior temporal atrophy for FTLD | ☐ 0 | ☐ 1 | ☐ 8 |
| 6j. Dopamine transporter scan (DATscan) evidence for Lewy body disease | ☐ 0 | ☐ 1 | ☐ 8 |
| 6k. Other (SPECIFY): ______ | ☐ 0 | ☐ 1 | |

**7. Is there evidence for cerebrovascular disease (CVD) on imaging?**

| Imaging findings | No | Yes | Unknown/ not assessed |
|---|---|---|---|
| 7a. Large vessel infarct(s) | ☐ 0 | ☐ 1 | ☐ 8 |
| 7b. Lacunar infarct(s) | ☐ 0 | ☐ 1 | ☐ 8 |
| 7c. Macrohemorrhage(s) | ☐ 0 | ☐ 1 | ☐ 8 |
| 7d. Microhemorrhage(s) | ☐ 0 | ☐ 1 | ☐ 8 |
| 7e. Moderate white-matter hyperintensity (CHS score 5–6) | ☐ 0 | ☐ 1 | ☐ 8 |
| 7f. Extensive white-matter hyperintensity (CHS score 7–8+) | ☐ 0 | ☐ 1 | ☐ 8 |

**8. Does the subject have a dominantly inherited AD mutation (PSEN1, PSEN2, APP)?**
☐ 0 No ☐ 1 Yes ☐ 9 Unknown/not assessed

**9. Does the subject have a hereditary FTLD mutation (e.g., GRN, VCP, TARBP, FUS, C9orf72, CHMP2B, MAPT)?**
☐ 0 No ☐ 1 Yes ☐ 9 Unknown/not assessed

**10. Does the subject have a hereditary mutation other than an AD or FTLD mutation?**
☐ 0 No ☐ 1 Yes (SPECIFY): ______ ☐ 9 Unknown/not assessed

**SECTION 3: Etiologic diagnoses**

Section 3 must be filled out for all subjects. Indicate presumptive etiologic diagnoses of the cognitive disorder and whether a given diagnosis is a primary, contributing, or non-contributing cause of the observed impairment, based on the clinician's best judgment. **Select one or more diagnoses as Present; all others will default to Absent in the NACC database.** Only one diagnosis should be selected as 1=Primary.

**For subjects with normal cognition:** Indicate the presence of any diagnoses by marking Present, and leave the questions on whether the diagnosis was primary, contributing, or non-contributing blank. Subjects with positive biomarkers but no clinical symptoms of Alzheimer's disease, Lewy body disease, or frontotemporal lobar degeneration **should not** have these diagnoses marked as Present. Instead, the biomarker data from Section 2 can be used to identify the presence of preclinical disease.

| Etiologic diagnoses | Present | Primary | Contributing | Non-contributing |
|---|---|---|---|---|
| 11. Alzheimer's disease | ☐ 1 | 11a ☐ 1 | ☐ 2 | ☐ 3 |
| 12. Lewy body disease<br>12b. ☐ 1 Parkinson's disease | ☐ 1 | 12a ☐ 1 | ☐ 2 | ☐ 3 |
| 13. Multiple system atrophy | ☐ 1 | 13a ☐ 1 | ☐ 2 | ☐ 3 |
| 14. Frontotemporal lobar degeneration | | | | |
| 14a. Progressive supranuclear palsy (PSP) | ☐ 1 | 14a1 ☐ 1 | ☐ 2 | ☐ 3 |
| 14b. Corticobasal degeneration (CBD) | ☐ 1 | 14b1 ☐ 1 | ☐ 2 | ☐ 3 |
| 14c. FTLD with motor neuron disease | ☐ 1 | 14c1 ☐ 1 | ☐ 2 | ☐ 3 |
| 14d. FTLD NOS | ☐ 1 | 14d1 ☐ 1 | ☐ 2 | ☐ 3 |
| 14e. If FTLD (Questions 14a – 14d) is Present, specify FTLD subtype:<br>☐ 1 Tauopathy<br>☐ 2 TDP-43 proteinopathy<br>☐ 3 Other (SPECIFY): ______<br>☐ 9 Unknown | | | | |

#### SECTION 3: Etiologic diagnoses (cont.)

Section 3 must be filled out for all subjects. Indicate presumptive etiologic diagnoses of the cognitive disorder and whether a given diagnosis is a primary, contributing, or non-contributing cause of the observed impairment, based on the clinician's best judgment. **Select one or more diagnoses as Present; all others will default to Absent in the NACC database.** Only one diagnosis should be selected as **1=Primary**.

**For subjects with normal cognition:** Indicate the presence of any diagnoses by selecting **1=Present**, and leave the questions on whether the diagnosis was primary, contributing, or non-contributing blank. Subjects with positive biomarkers but no clinical symptoms of Alzheimer's disease, Lewy body disease, or frontotemporal lobar degeneration **should not** have these diagnoses selected as Present. Instead, the biomarker data from Section 2 can be used to identify the presence of preclinical disease.

| Etiologic diagnoses | Present | Primary | Contributing | Non-contributing |
|---|---|---|---|---|
| 15. Vascular brain injury (based on clinical or imaging evidence)<br>*If significant vascular brain injury is absent,* **SKIP TO QUESTION 16.**<br>15b. Previous symptomatic stroke?<br>☐0 No **(SKIP TO QUESTION 15c)**<br>☐1 Yes<br>15b1. Temporal relationship between stroke and cognitive decline?<br>☐0 No<br>☐1 Yes<br>15b2. Confirmation of stroke by neuroimaging?<br>☐0 No<br>☐1 Yes<br>☐9 Unknown; no relevant imaging data available<br>15c. Is there imaging evidence of cystic infarction in cognitive network(s)?<br>☐0 No<br>☐1 Yes<br>☐9 Unknown; no relevant imaging data available<br>15d. Is there imaging evidence of cystic infarction, imaging evidence of extensive white matter hyperintensity (CHS grade 7–8+), <u>and</u> impairment in executive function?<br>☐0 No<br>☐1 Yes<br>☐9 Unknown; no relevant imaging data available | ☐1 | 15a ☐1 | ☐2 | ☐3 |
| 16. Essential tremor | ☐1 | 16a ☐1 | ☐2 | ☐3 |
| 17. Down syndrome | ☐1 | 17a ☐1 | ☐2 | ☐3 |
| 18. Huntington's disease | ☐1 | 18a ☐1 | ☐2 | ☐3 |
| 19. Prion disease (CJD, other) | ☐1 | 19a ☐1 | ☐2 | ☐3 |

| Etiologic diagnoses | Present | Primary | Contributing | Non-contributing |
|---|---|---|---|---|
| 20. Traumatic brain injury<br>20b. If Present, does the subject have symptoms consistent with chronic traumatic encephalopathy?<br>☐0 No ☐1 Yes ☐9 Unknown | ☐1 | 20a ☐1 | ☐2 | ☐3 |
| 21. Normal-pressure hydrocephalus | ☐1 | 21a ☐1 | ☐2 | ☐3 |
| 22. Epilepsy | ☐1 | 22a ☐1 | ☐2 | ☐3 |
| 23. CNS neoplasm<br>23b. ☐1 Benign ☐2 Malignant | ☐1 | 23a ☐1 | ☐2 | ☐3 |
| 24. Human immunodeficiency virus (HIV) | ☐1 | 24a ☐1 | ☐2 | ☐3 |
| 25. Cognitive impairment due to other neurologic, genetic, or infectious conditions not listed above<br>25b. If Present, specify: ______________________ | ☐1 | 25a ☐1 | ☐2 | ☐3 |

Section 3 must be filled out for all subjects. Indicate presumptive etiologic diagnoses of the cognitive disorder and whether a given diagnosis is a primary, contributing, or non-contributing cause of the observed impairment, based on the clinician's best judgment. **Select one or more diagnoses as Present; all others will default to Absent in the NACC database.** Only one diagnosis should be selected as **1=Primary.**

**For subjects with normal cognition:** Indicate the presence of any diagnoses by selecting **1=Present**, and leave the questions on whether the diagnosis was primary, contributing, or non-contributing blank. Subjects with positive biomarkers but no clinical symptoms of Alzheimer's disease, Lewy body disease, or frontotemporal lobar degeneration **should not** have these diagnoses selected as Present. Instead, the biomarker data from Section 2 can be used to identify the presence of preclinical disease.

| Condition | Present | Primary | Contributing | Non-contributing |
|---|---|---|---|---|
| 26. Active depression<br>26b. If Present, select one:<br>☐0 Untreated<br>☐1 Treated with medication and/or counseling | ☐1 | 26a ☐1 | ☐2 | ☐3 |
| 27. Bipolar disorder | ☐1 | 27a ☐1 | ☐2 | ☐3 |
| 28. Schizophrenia or other psychosis | ☐1 | 28a ☐1 | ☐2 | ☐3 |
| 29. Anxiety disorder | ☐1 | 29a ☐1 | ☐2 | ☐3 |
| 30. Delirium | ☐1 | 30a ☐1 | ☐2 | ☐3 |
| 31. Post-traumatic stress disorder (PTSD) | ☐1 | 31a ☐1 | ☐2 | ☐3 |
| 32. Other psychiatric disease<br>32b. If Present, specify: ______________________ | ☐1 | 32a ☐1 | ☐2 | ☐3 |

| | | | | |
|---|---|---|---|---|
| 33. Cognitive impairment due to alcohol abuse<br>33b. Current alcohol abuse:<br>☐0 No ☐1 Yes ☐9 Unknown | ☐1 | 33a ☐1 | ☐2 | ☐3 |
| 34. Cognitive impairment due to other substance abuse | ☐1 | 34a ☐1 | ☐2 | ☐3 |
| 35. Cognitive impairment due to systemic disease/ medical illness (as indicated on Form D2) | ☐1 | 35a ☐1 | ☐2 | ☐3 |
| 36. Cognitive impairment due to medications | ☐1 | 36a ☐1 | ☐2 | ☐3 |
| 37. Cognitive impairment NOS<br>37b. If Present, specify: ______________________ | ☐1 | 37a ☐1 | ☐2 | ☐3 |
| 38. Cognitive impairment NOS<br>38b. If Present, specify: ______________________ | ☐1 | 38a ☐1 | ☐2 | ☐3 |
| 39. Cognitive impairment NOS<br>39b. If Present, specify: ______________________ | ☐1 | 39a ☐1 | ☐2 | ☐3 |

**sTable 2 Demographic and clinical characteristics of participants after propensity score matching for diclofenac vs no NSAID use.**

Median (Interquartile Range (IQR); n (%).

| **Variable** | **Diclofenac** N = 702 | **No NSAID** N = 702 | **P-value** |
|---|---|---|---|
| **Age (years)** | 77 (71-84) | 77 (71-84) | 0.9 |
| **Sex** | | | 0.8 |
| Male | 206 (29%) | 200 (28%) | |
| Female | 496 (71%) | 502 (72%) | |
| **Race** | | | >0.9 |
| White | 537 (76%) | 542 (77%) | |
| Black or African American | 114 (16%) | 115 (16%) | |
| American Indian or Alaska Native | 5 (0.7%) | 3 (0.4%) | |
| Native Hawaiian or Pacific Islander | 1 (0.1%) | 1 (0.1%) | |
| Asian | 12 (1.7%) | 9 (1.3%) | |
| Multiracial | 33 (4.7%) | 32 (4.6%) | |
| **Education** (years) | 16 (13-18) | 16 (13-18) | >0.9 |
| **MoCA-Score** (Education-corrected) | 25 (21-28) | 24 (20-27) | **0.033** |
| **Cognitive Status** (demented) | 201 (29%) | 246 (35%) | **0.012** |
| **Evidence of Alzheimer’s disease** | | | **0.005** |
| Normal cognition | 397 (57%) | 343 (49%) | |
| Diagnosis (presumptive) | 305 (43%) | 359 (51%) | |
| No Diagnosis | 0 (0%) | 0 (0%) | |
| **Elevated Tau in Cerebrospinal Fluid** | | | >0.9 |
| Negative | 14 (52%) | 12 (50%) | |

| Variable | Diclofenac<br>N = 702 | No NSAID<br>N = 702 | P-value |
|---|---|---|---|
| Positive | 13 (48%) | 12 (50%) | |
| **Low Amyloid Cerebrospinal Fluid** | | | 0.9 |
| Negative | 14 (52%) | 11 (46%) | |
| Positive | 13 (48%) | 13 (54%) | |
| **Cerebrospinal Fluid- Status** | | | 0.2 |
| Elevated Tau and low Amyloid | 9 (36%) | 11 (48%) | |
| Normal Tau and normal Amyloid | 10 (40%) | 11 (48%) | |
| Only low Amyloid | 3 (12%) | 1 (4.3%) | |
| Only elevated Tau | 3 (12%) | 0 (0%) | |

**sTable 3 Demographic and clinical characteristics of participants after propensity score matching for naproxen vs no NSAID use.**

Median (Interquartile Range (IQR); n (%).

| **Variable** | **Naproxen** n = 2,210 | **No NSAID** n = 2,210 | **P-value** |
|---|---|---|---|
| **Age (years)** | 76 (70-83) | 76 (70-83) | >0.9 |
| **Sex** | | | 0.6 |
| Male | 801 (36%) | 782 (35%) | |
| Female | 1,409 (64%) | 1,428 (65%) | |
| **Race** | | | 0.5 |
| White | 1,731 (78%) | 1,751 (79%) | |
| Black or African American | 329 (15%) | 300 (14%) | |
| American Indian or Alaska Native | 16 (0.7%) | 13 (0.6%) | |
| Native Hawaiian or Pacific Islander | 1 (<0.1%) | 4 (0.2%) | |
| Asian | 31 (1.4%) | 28 (1.3%) | |
| Multiracial | 102 (4.6%) | 114 (5.2%) | |
| **Education (years)** | 16 (13-18) | 16 (13-18) | 0.6 |
| **MoCA-Score (Education-corrected)** | 25 (21-28) | 25 (19-27) | **<0.001** |
| **Cognitive Status (demented)** | 689 (31%) | 852 (39%) | **<0.001** |
| **Evidence of Alzheimer's disease** | | | **<0.001** |
| Normal cognition | 1,221 (55%) | 1,065 (48%) | |
| Diagnosis (presumptive) | 989 (45%) | 1,145 (52%) | |
| No Diagnosis | 0 (0%) | 0 (0%) | |

| Variable | Naproxen<br>n = 2,210 | No NSAID<br>n = 2,210 | P-value |
|---|---|---|---|
| **Elevated Tau in Cerebrospinal Fluid** | | | 0.14 |
| Negative | 43 (58%) | 33 (45%) | |
| Positive | 31 (42%) | 41 (55%) | |
| **Low Amyloid Cerebrospinal Fluid** | | | 0.057 |
| Negative | 42 (53%) | 27 (36%) | |
| Positive | 38 (48%) | 48 (64%) | |
| **Cerebrospinal Fluid- Status** | | | 0.12 |
| Elevated Tau and low Amyloid | 25 (34%) | 37 (53%) | |
| Normal Tau and normal Amyloid | 32 (43%) | 24 (34%) | |
| Only low Amyloid | 11 (15%) | 6 (8.6%) | |
| Only elevated Tau | 6 (8.1%) | 3 (4.3%) | |

**sTable 4 Demographic and clinical characteristics of participants after propensity score matching for etodolac vs no NSAID use.**

Median (Interquartile Range (IQR); n (%).

| Variable | Etodolac N = 125 | No NSAID N = 125 | P-value |
|---|---|---|---|
| **Age (years)** | 78 (71-85) | 78 (71-85) | >0.9 |
| **Sex** | | | 0.5 |
| Male | 53 (42%) | 47 (38%) | |
| Female | 72 (58%) | 78 (62%) | |
| **Race** | | | 0.7 |
| White | 107 (86%) | 110 (88%) | |
| Black or African American | 13 (10%) | 12 (9.6%) | |
| American Indian or Alaska Native | 1 (0.8%) | 0 (0%) | |
| Native Hawaiian or Pacific Islander | 0 (0%) | 0 (0%) | |
| Asian | 0 (0%) | 0 (0%) | |
| Multiracial | 4 (3.2%) | 3 (2.4%) | |
| **Education (years)** | 16 (12-18) | 16 (12-18) | 0.6 |
| **MoCA score (Education-corrected)** | 25 (20-27) | 25 (17-28) | 0.6 |
| **Cognitive Status** (demented) | 46 (37%) | 52 (42%) | 0.5 |
| **Evidence of Alzheimer's disease** | | | 0.4 |
| Normal cognition | 63 (50%) | 55 (44%) | |
| Diagnosis (presumptive) | 62 (50%) | 70 (56%) | |
| **Elevated Tau in Cerebrospinal Fluid** | | | >0.9 |

| Variable | Etodolac<br>N = 125 | No NSAID<br>N = 125 | P-value |
|---|---|---|---|
| Negative | 0 (0%) | 1 (25%) | |
| Positive | 2 (100%) | 3 (75%) | |
| **Low Amyloid Cerebrospinal Fluid** | | | |
| Negative | 2 (100%) | 5 (100%) | |
| Positive | 123 | 120 | |
| **Cerebrospinal Fluid- Status** | | | >0.9 |
| Elevated Tau and low Amyloid | 2 (100%) | 3 (75%) | |
| Only low Amyloid | 0 (0%) | 1 (25%) | |

**sTable 5 Selected study population from UDS versions 3.0 and 3.2.**

Median (Interquartile Range (IQR); n (%).

| Variable | Naproxen n = 1,328 | Diclofenac n = 567 | Etodolac n = 43 | No NSAID n = 22,273 | P-value |
|---|---|---|---|---|---|
| **Age** (years) | 75 (69-81) | 76 (70-83) | 73 (68-78) | 75 (68-82) | **0.002** |
| **Sex** | | | | | **<0.001** |
| Male | 520 (39%) | 186 (33%) | 18 (42%) | 9,480 (43%) | |
| Female | 808 (61%) | 381 (67%) | 25 (58%) | 12,793 (57%) | |
| **Race** | | | | | **<0.001** |
| White | 1,045 (79%) | 409 (72%) | 35 (81%) | 17,724 (80%) | |
| Black or African American | 181 (14%) | 102 (18%) | 6 (14%) | 2,734 (12%) | |
| American Indian or Alaska Native | 7 (0.5%) | 6 (1.1%) | 0 (0%) | 125 (0.6%) | |
| Native Hawaiian or Pacific Islander | 2 (0.2%) | 0 (0%) | 0 (0%) | 18 (<0.1%) | |
| Asian | 25 (1.9%) | 12 (2.1%) | 0 (0%) | 726 (3.3%) | |
| Multiracial | 59 (4.4%) | 29 (5.1%) | 1 (2.3%) | 579 (2.6%) | |
| Unknown | 9 (0.7%) | 9 (1.6%) | 1 (2.3%) | 367 (1.6%) | |
| **Education** (years) | 16 (14-18) | 16 (14-18) | 16 (14-18) | 16 (14-18) | 0.6 |
| **MoCA score** | 25 (21- 28) | 25 (20-27) | 25 (20-27) | 24 (18-27) | **<0.001** |

| Variable | Naproxen<br>n = 1,328 | Diclofenac<br>n = 567 | Etodolac<br>n = 43 | No NSAID<br>n = 22,273 | P-value |
|---|---|---|---|---|---|
| (Education-corrected) | | | | | |
| **Cognitive Status** (demented) | 334 (25%) | 139 (25%) | 10 (23%) | 7,533 (34%) | **<0.001** |
| **Evidence of Alzheimer's disease** | | | | | **<0.001** |
| Normal cognition | 202 (15%) | 97 (17%) | 4 (9.3%) | 4,069 (18%) | |
| Diagnosis (presumptive) | 411 (31%) | 178 (31%) | 16 (37%) | 8,435 (38%) | |
| No Diagnosis | 715 (54%) | 292 (51%) | 23 (53%) | 9,769 (44%) | |
| **Elevated Tau in Cerebrospinal Fluid** | | | | | 0.2 |
| Negative | 42 (58%) | 17 (57%) | 0 (0%) | 702 (53%) | |
| Positive | 30 (42%) | 13 (43%) | 2 (100%) | 621 (47%) | |
